# Long-read sequencing reveals two common *APOE* ε3 and ε4 intragenic haplotypes in the Spanish population

**DOI:** 10.1101/2025.03.25.25324541

**Authors:** Pablo García-González, Raquel Puerta, Álvaro Muñoz-Morales, Claudia Olivé, Berta Calm, Laura Montrreal, Alejandro Valenzuela, Paula Bayón-Buján, Marta Rovira, Maria Capdevila-Bayo, Marta Marquié, Sergi Valero, Maitee Rosende-Roca, Montserrat Alegret, Adrián Hinojosa, Pilar Sanz, Frederik Brosseron, Pamela Martino-Adami, Itziar de Rojas, Michael Heneka, Alfredo Ramirez, Arcadi Navarro, Maria E. Sáez, Lluís Tárraga, Jose E. Cavazos, Mercè Boada, Amanda Cano, Maria Victoria Fernández, Alfredo Cabrera-Socorro, Agustín Ruiz

## Abstract

The *APOE* gene is a key genetic determinant of Alzheimer’s disease (AD) risk. While the pathogenic effects of the ε4 allele are well established, the contribution of distinct haplotypic backgrounds within ε3 and ε4 remains poorly understood. Here, we used long-read Oxford Nanopore sequencing to identify and phase single nucleotide variants (SNVs) across a 3.9 kb region spanning *APOE* locus (3’ to 5’ UTR) in a large memory-clinic based cohort (*N=*1,266). We identified 48 SNVs, including nine novel variants, and reconstructed haplotypes through direct phasing. Our analysis revealed a structured haplotypic architecture, with two major configurations for ε3 (ε3a and ε3b) and ε4 (ε4a and ε4b), defined by the promoter variant rs405509 in phase with the canonical isoform-defining variants rs429358 and rs7412. The observed homoplasy in the rs405509, likely arising from a recombination or gene conversion event, complicates phasing by standard genetic approaches, highlighting the value of long-read sequencing for accurate characterization of *APOE* haplotypic diversity. Importantly, the ε4a haplogenotype was associated with reduced risk of progression from mild cognitive impairment to AD dementia compared to ε4b (*APOE*-stratified meta-analysis *HR[95%CI]=*0.564[0.400–0.794], *p=*0.001), while ε3b showed a borderline protective effect towards AD biomarker positivity in ε3ε3 subjects (*OR[95%CI]=*0.76[0.57–1.00], *p=*0.053). Notably, haplogenotypes had no detectable effect on CSF APOE levels, indicating that haplotype-dependent effects are not mediated by differences in total APOE abundance. This study provides new insights into the intragenic allelic variability of the *APOE* gene, highlighting the importance of considering haplotype-specific effects when interpreting the functional impact of *APOE* and in designing targeted therapeutic strategies. Further research is needed to explore the broader regulatory network of the *APOE* locus and its interaction with neighboring loci in the 19q13 region.

## 1 Introduction

Alzheimer’s disease (AD) and related dementias present a growing global health challenge as populations age, with significant social and economic burdens^1,2^. AD, the most common cause of dementia, is characterized by a progressive decline in cognitive function, leading to memory impairment, loss of independence, and ultimately, death. The profound effects on patients and caregivers, coupled with the strain on healthcare systems, highlight the urgent need for effective strategies to prevent and manage this disease^3^. Genetic factors are critical in developing sporadic, late-onset AD (LOAD), with the *APOE* gene identified as the strongest genetic risk factor. Specifically, the *APOE* ε4 allele significantly increases the risk for AD, while the ε2 allele shows a protective effect^4,5^. However, the risk associated with the *APOE* ε4 allele varies across populations^6–11^. Notably, APOE-related AD risk is markedly reduced in individuals of African ancestry compared to those of European descent, suggesting the presence of protective effects within specific genetic backgrounds^12^. Recent studies have highlighted a significant interaction between *APOE* ε4 and the rs10423769-A allele, which reduces AD risk by up to 75% in African-origin haplotypes^12^. Understanding the genetic mechanisms modulating *APOE* and its association with AD—particularly the role of cis and trans regulatory elements—remains critical for developing targeted interventions aimed at mitigating the impact of this major risk factor and ultimately reducing the societal burden of the disease.

The *APOE* gene encodes a major regulator of lipid metabolism binding and transporting lipids within the bloodstream and the brain. In neurodegenerative diseases, *APOE* plays pivotal roles in amyloid-β clearance, synaptic maintenance, and neuroinflammation^13,14^. The *APOE* ε4 isoform has been shown to impair amyloid-β clearance, promoting its accumulation in the brain, a hallmark of AD pathology^15^. Furthermore, the ε4 isoform is associated with enhanced neuroinflammation^16^ and oxidative stress^17^, contributing to neuronal damage and accelerating disease progression. These distinct roles of *APOE* isoforms emphasize the importance of understanding their functional differences, particularly in the context of AD pathology and progression^18^.

Homoplasy, an event in which a same genetic feature arises independently in multiple evolutionary events, is a well-known issue within the *APOE* locus, complicating efforts to link genetic variants with disease phenotypes^19^. Long-read sequencing technology provides the ability to phase DNA variations, enabling the precise connection of single nucleotide variants (SNVs) with haplotypes and regulatory elements that modulate *APOE* gene expression^20,21^. This level of resolution facilitates a clearer distinction between benign and disease-associated variants, enhancing our understanding of how different *APOE* haplotypes influence disease risk and progression across diverse populations^22^. The ability to phase the classic *APOE* isoforms with other variants located in functional elements provides an invaluable tool for elucidating the mechanisms by which cis-genetic variants configure *APOE* sub-haplotypes. This understanding extends beyond the classical ε2/ε3/ε4 isoforms, shedding light on how specific combinations contribute to Alzheimer’s disease and other neurodegenerative conditions

In this study, we explore the association between intragenic *APOE* haplotypes and AD pathogenesis, with a special focus on identifying existing haplotypes within the ε3 and ε4 protein isoforms. Utilizing Oxford Nanopore Technology (ONT) based long-read sequencing, we analyzed the distribution of intragenic *APOE* sub-haplotypes in a large Spanish cohort (*N=*1,266), identifying 48 SNVs, including both known and novel variants. Our analysis specifically focuses on examining common variants within regulatory motifs that influence CSF APOE levels and AD biomarkers in an isoform-specific manner. Moreover, we identified distinct ε3 and ε4 sub-haplotypes that appear to influence the rate of conversion from mild cognitive impairment (MCI) to dementia, providing potential genetic markers for individualized disease risk assessment and therapeutic intervention.

## 2 Materials and methods

### 2.1 The ACE cohort

Patient recruitment and evaluation were conducted at the Memory Disorders Unit of ACE Alzheimer Center Barcelona (ACE) in Spain, spanning the years 2016 to 2021^23^. Diagnoses were established through consensus discussions involving neurologists, neuropsychologists, and social workers during case conferences. All patients classified as having mild cognitive impairment (MCI) met the criteria outlined by *Petersen et al.* for MCI diagnosis^24,25^. These criteria include subjective memory complaints, a noticeable decline from normal cognitive function, intact daily living activities, the absence of dementia, and measurable impairments in one or more cognitive domains, whether amnestic MCI (single or multiple domains). Impairment thresholds were determined according to age and education level. Specific cutoffs for the tests in the neuropsychological battery (NBACE) are detailed in prior publications^26^, with individuals scoring below these thresholds classified as having MCI. For patients with MCI who were later followed up, dementia diagnoses were made according to the DSM-V criteria^27^. Dementia subtypes were further categorized based on the 2011 NIA-AA criteria for Alzheimer’s disease^28^, the NINDS-AIREN criteria for vascular dementia^29^, frontotemporal dementia^30^, and the diagnostic criteria for Lewy body dementia^31^.

Fasting patients provided paired CSF and plasma samples using standardized clinical methods. CSF was collected via lumbar puncture from the L3-L4 intervertebral space following standard protocols^32^. The procedure, performed by skilled neurologists under local anesthesia (1% mepivacaine), involved the patient sitting upright. Two 10-ml polypropylene tubes (Sarstedt ref 62,610,018) of CSF were collected, with one tube used for routine biochemistry tests (glucose, total protein, proteinogram, cell type, and count). The second tube was centrifuged (2000xg, 10 min at 4°C), aliquoted into polypropylene tubes (Sarstedt ref 72,694,007), and stored at –80°C within two hours of collection. On the day of biomarker analysis, an aliquot was thawed at room temperature, vortexed for 5–10 seconds, and analyzed for amyloid-β 1-42 (Aβ42), total tau (t-tau), and 181-phosphorylated tau (p-tau) levels using enzyme-linked immunosorbent assays (ELISAs): Innotest Aβ1-42, Innotest hTAU Ag, and Innotest PHOSPHO-TAU (181P) (Fujirebio Europe)^32–34^.

### 2.2 DNA extraction and *APOE* genotyping

DNA extraction from blood specimens was performed automatically using standard procedures with the DNA Chemagic system (Perkin Elmer) or the Maxwell RSC48 instrument (Promega). Extensive DNA quality control was conducted, and only samples with DNA concentrations greater than 10 ng/µl and high integrity were included for *APOE* genotyping. The isoforms were determined by TaqMan probe analysis in the Real-Time PCR QuantStudio3 (ThermoFisher, Waltham, Massachusetts, USA) system or extracted from Affymetrix Axiom SP biobank arrays processed as previously described^35–37^.

### 2.3 Long PCR amplification of the *APOE* locus

Long-range PCR was performed to amplify the *APOE* locus using primer pairs designed with the National Institutes of Health (NIH) primer designing tool software (https://www.ncbi.nlm.nih.gov/tools/primer-blast/) to cover the entire region, from the promoter to the 3’ UTR. We used Roche’s Expand™ Long Template PCR System kit (UNSPSC number: 41106300) for PCR amplification. To find the optimal reaction conditions, we tested in a single sample three different primer pairs, along with different DMSO concentrations (2.5, 5.0, 7.5 and 10%) and three kit-supplied reaction buffers (Supp. Table 1, Supp. Fig. 1). Based on our results, we used a 10% concentration of DMSO, kit-supplied reaction buffer 2 and the primer pair labeled “APOE_III” for optimal long-range amplification of the region. The selected primers were: forward primer 5’-ACAGGGTCAGGAAAGGAGGA –3’ and reverse primer 5’-GGCTGGGGCTTAGAGGAAAT –3’, spanning a 3939 bp fragment surrounding the *APOE* locus. The reaction mixture included a high-fidelity DNA polymerase, PCR dNTP mix (Roche, UNSPSC code 41106300), template DNA, primers, DMSO, and buffer (including 17.5 mM MgCl₂) (Supp. Table 2). The PCR was carried out on a Thermo Fisher QuantStudio 3 Real-Time Thermal Cycler with the following conditions: initial denaturation at 94°C for 2 minutes, followed by 40 cycles of denaturation at 92°C for 10 seconds, annealing at 59°C for 30 seconds, and extension at 68°C for 4 minutes. A ramp increasing the elongation time by 20 seconds per cycle starting in cycle 11, and a final 10-minute elongation step at 68°C were included in the protocol to ensure complete amplification of the DNA segment. Following amplification, PCR products were purified using AMpure XP beads (Beckman Coulter, A63881), following the manufacturer’s protocol to remove salts, primers, nucleotides, and enzymes, ensuring high-quality DNA for downstream applications. Amplification success was verified via automated capillary electrophoresis using the LabChip GX Touch Nucleic Acid Analyzer (Revvity, CLS138162) and the DNA 12K Reagent Kit (Revvity, 760569).

### 2.4 ONT long-read Sequencing and Library Preparation

For the ONT sequencing of the *APOE* amplicon, we employed the ONT 96 Native Barcoding Kit (EXP-NBD196) to multiplex 96 samples in a single reaction, and the ONT Ligation Sequencing Kit (SQK-LSK109). We followed the protocol recommended by ONT based on these reagent kits: “Amplicon barcoding with Native Barcoding Expansion 96 (EXP-NBD196, and SQK-LSK109), Version: NBA_9102_v109_revA_09Jul2020”. Purified *APOE* amplicons (100 fmoles) were quantified using the Quant-iT™ PicoGreen™ dsDNA Assay Kits (Invitrogen, P7589), and 250 ng were taken forward for barcode ligation. This process involved two steps: first, the ends of the DNA fragments were repaired using the NEBNext® Ultra™ II End Repair/dA-Tailing Module (New England Biolabs, E7546), following manufacturer instructions. This reaction is optimized for the subsequent ligation reaction, yielding 5’ phosphorylated and 3’ dA-tailed ends. Then, unique barcodes were ligated to each sample using the NEB Blunt/TA Ligase Master Mix module (New England Biolabs, M0367). Subsequently, the barcoded library was pooled together and 480 uL were carried forward for purification using Ampure XP beads, removing excess barcodes and small fragments, and collected in a volume of 30 uL. Absorbance at 260 nm, A260/A280 and A260/230 ratios, measured with a BioTek Epoch Microplate Spectrophotometer, were used subsequently for rapid estimation of DNA concentration and purity. 500 ng (200 fmol) of the pulled library was then ligated to ONT sequencing adapters using the NEBNext Quick Ligation Module (New England Biolabs, E6056), following manufacturer instructions. A final AMpure XP purification step was performed to remove unwanted products. Finally, 100 ng (∼40 fmoles) of the library were loaded onto primed ONT R9.4.1 flow cells. Sequencing was carried out on a MinION platform, enabling real-time demultiplexing by Guppy, until satisfactory coverage of all barcoded samples was achieved (at least 100x coverage of the least represented barcodes).

### 2.5 ONT long-read variant calling

The dry lab workflow for the identification of SNVs in the *APOE* locus using ONT sequencing data is based on the protocol described by *Leija-Salazar et al.*^38^. This bioinformatics processing involves a series of computational steps, starting from raw signal data and culminating in the generation of a multi-sample VCF file. The process begins with basecalling and demultiplexing using Guppy v4.5.2 (https://nanoporetech.com/software/other/guppy), which converts raw electrical signals from the nanopore sequencing (fast5) into nucleotide sequences (fastq files), while simultaneously assigning each read to its corresponding barcode. Basecalling was conducted using the dna_r9.4.1_450bps_hac.cfg config file for high accuracy. We focused exclusively on SNVs due to the known limitations of ONT sequencing, which tends to be less reliable for detecting indel mutations^39^. By limiting our analysis to SNVs, we leverage the strengths of ONT sequencing, ensuring higher confidence in variant detection and reducing the risk of false positives typically associated with indel identification. We generated in-silico gel electrophoresis images to visually assess the read length of sequencing reads, ensuring the signal detected for each barcode was present in the expected length (3939bp). Following this, the quality of the reads is assessed; only those marked as “PASS” and within the length range of 3800 to 4000 base pairs were retained to ensure only full-length *APOE* amplicons were included in downstream analysis. Subsequently, *nanopolish* (https://github.com/jts/nanopolish) was utilized to index the fastq files, linking the sequencing data back to the original raw signal data in the fast5 files. This linkage is crucial for the later implementation of signal-based error correction during variant calling. We used two different aligners—*Graphmap* v0.5.2 and *NGLMR* v0.2.7—to generate the SAM files providing the positional information of the filtered and indexed reads within the reference genome (GRCh38.p13). Subsequently, we used *samtools* v1.10 (http://www.htslib.org/) to convert the files to BAM format, and then sort and index the files. nanopolish was subsequently used for variant calling, employing the raw nanopore signal to enhance the accuracy of mutation detection. The output of this step is a vcf file containing the identified variants within the *APOE* locus for each sample. Individual vcf files from each sample were then merged using *bcftools* 1.9 (https://samtools.github.io/bcftools), and sample IDs were reassigned based on barcode correspondence, resulting in a multi-sample vcf file that consolidates the variant information across all 96 samples. SNP identifiers and TOPMed allele frequencies were annotated based on dbSNP build 151 (GRCh38.p7). The mean ratio of the QUAL and TotalReads parameters generated by nanopolish was used as a quality measure for variant calling. Overall, this comprehensive approach allowed for the accurate detection of missense mutations in the *APOE* locus by leveraging both basecalled sequences and the inherent error-correction capabilities of Nanopolish based on the raw nanopore sequencing data. Furthermore, variant functional annotation was performed using Ensembl’s Variant Effect Predictor (VEP) tool^40^. ClinVar clinical significance annotations were collapsed into a single category per variant using a custom R function. Multi-valued annotations were parsed and prioritized such that conflicting or mixed pathogenic/benign labels were classified as VUS, while remaining variants were assigned to pathogenic/likely pathogenic, benign/likely benign, or retained as non-Mendelian categories (e.g., risk factor, protective, drug response).

### 2.6 Variant phasing and further haplotype analyses

The phasing of *APOE* haplotypes was conducted using *WhatsHap* (https://whatshap.readthedocs.io), a tool designed for accurate read-based phasing of long-read sequencing data^41,42^. *WhatsHap* performs phasing by directly utilizing DNA sequencing reads, a method known as read-based phasing or haplotype assembly, which is particularly suited for long reads but also works well with short reads. By leveraging the long-range information from ONT sequencing, *WhatsHap* effectively separated detected variants into their respective haplotypes, enabling the reconstruction of complete *APOE* haplotypes for each individual. To facilitate downstream analyses, we developed a custom code. This program automated the extraction of individual-level haplotypes from the phased VCFs, calculated haplotype frequencies, and encoded individual-level subhaplotypes for subsequent use.

Following the phasing process, the evolutionary relationships of the *APOE* haplotypes were examined using MEGA Software v11.0.13 (https://www.megasoftware.net)^43^. MEGA was used for the alignment and comparison of haplotype sequences, enabling the construction of phylogenetic trees and the assessment of evolutionary divergence. Specifically, the evolutionary history was inferred using the Maximum Likelihood method with the Tamura-Nei model^44^. The tree was drawn to scale, with branch lengths representing the number of substitutions per site. Sub-haplotypes derived from the different *APOE* common haplotypes were color-coded to facilitate interpretation: green for ε2, orange for ε3, red for ε4, and blue for the ancestral sequence extracted from Ensembl^45^.

### 2.7 CSF APOE protein measurements

CSF APOE levels measured using the Olink platform were obtained from the Inflamation II panel generated in our previous work as part of the PREADAPT JPND program^46^. Following standard procedures, values below the limit of detection were removed and protein values were log2-transformed. Samples without available Olink measurements were analyzed using the Lumipulse platform (N=744) according to manufacturer instructions. This assay enables simultaneous quantification of total APOE (Pan-APOE; REF: 81449, Fujirebio Europe, Belgium) and the APOE4 isoform (REF: 81453, Fujirebio Europe, Belgium). APOE4 measurements were restricted to samples carrying at least one *APOE* ε4 allele (N=268). Based on manufacturer guidelines, an APOE4/Pan-APOE ratio of 5-75% indicates *APOE* ε4 heterozigosity, while a ratio of >=75% indicates ε4 homozigosity. Samples with discordant *APOE* genotype and proteotype (N=6) were excluded from further analysis. Pan-APOE protein levels were log-transformed prior to statistical analysis.

### 2.8 Statistical analysis

We employed R v4.1.1. to conduct statistical analyses and data processing. Baseline was defined as the clinical assessment closest in time to lumbar puncture. All reported p-values are two-sided. To evaluate the effect of *APOE* haplogenotypes on progression from MCI to AD dementia, Kaplan–Meier curves were generated using the *survminer* package. Differences in survival curves, reflecting variation in progression rates, were assessed using the log-rank test implemented in the *ggsurvplot* function. Multivariate models testing the association of *APOE* haplogenotypes with AD clinical features and endophenotypes were described within Supp. Table 3. Differences on progression risk were assessed by Cox proportional hazards models adjusted by age, sex and baseline MMSE using the *survival* R package^47^. Associations with AD pathology were assessed using logistic regression models comparing A+T+ and A-T-individuals, adjusted for age at baseline and sex. Logistic regression model performance was evaluated using Nagelkerke’s pseudo-R^2^, a scaled version of the Cox & Snell R^2^ statistic^48,49^. To evaluate the effect of haplogenotypes on CSF APOE protein levels, we fitted linear models adjusting for inter-individual variability in CSF turnover and blood–brain barrier integrity^50^, using OPCML levels and the second principal component derived from SomaScan CSF proteomics data as covariates. Adjusted CSF APOE levels were obtained as the residuals derived from linear regressions fitting APOE levels to OPCML (OBCAM_seq.15622.13 somamer) and the second principal component derived from Somascan CSF proteomics data. Analyses were performed separately for Olink and Lumipulse datasets. Pairwise comparisons of APOE protein levels between haplogenotypes were performed using t-tests.

## 3 Results

### 3.1 ACE cohort description

A detailed overview of the demographic, clinical, neurological, and biological characteristics of the ACE cohort is provided (Table 1). The ACE cohort is predominantly female (57.7%), with a mean age of 72.7 years, a mean body mass index (BMI) of 26.9 kg/m², and a mean educational attainment of 8.2 years. Based on Clinical Dementia Rating (CDR) scores, 7.2% of participants were cognitively unimpaired (CU; CDR=0), 62.7% had mild cognitive impairment (MCI, CDR = 0.5), and 30.1% had dementia (CDR >= 1) at baseline. Cognitive performance, as measured by the Mini-Mental State Examination (MMSE), had a mean score of 24.4. AT classification^51^ based on CSF Aβ42 and p-tau181 levels showed that 38.3% of individuals were A+T+, indicating concomitant amyloid and tau pathology, whereas 30.7% were A-T-, with no evidence of AD pathology. The remaining 31.0% displayed intermediate AT profiles (A+T– or A-T+), consistent with partial or evolving pathological changes that do not meet the full molecular criteria for AD. *APOE* genotype distribution was consistent with expectations, with ε3/ε3 being the most frequent genotype (57.9%), followed by ε3/ε4 (27.9%) and lower frequencies for ε2/ε3 (7.2%), ε4/ε4 (4.7%), ε2/ε4 (1.9%), and ε2/ε2 (0.3%). As expected, the prevalence of APOE ε4 alleles and CSF AT biomarker positivity increased with higher CDR categories, whereas APOE ε2 frequency decreased. Among the MCI individuals with available follow-up (N=724), 45.3% progressed to AD dementia, 8.7% to non-AD dementia, and 46.0% remained stable (Supp. Table 4). Baseline MMSE, age and *APOE* ε4 genotype were strongly associated with clinical progression to AD dementia (Supp. Table 4).

**Table 1.** ACE cohort demographics split by disease status: cognitively unimpaired (CU), mild cognitive impairment (MCI) and dementia. APOE genotypes were previously determined using Thermo Fisher qPCR TaqMan probes targeting rs429358 and rs7412, and/or genome-wide genotyping arrays (see methods); p-tau and Aβ42 levels are expressed in pg/mL units. ELISA values were transformed to their Lumipulse equivalents based on previously determined Passing-Bablok coefficients^34^.

|  | **CU**<br>**(CDR=0)** | **MCI**<br>**(CDR=0.5)** | **Dementia**<br>**(CDR>=1)** | **All** |
| --- | --- | --- | --- | --- |
| Sample Size | 91 (7.2%) | 794 (62.7%) | 381 (30.1%) | 1266 |
| Age (Mean, SD) | 63.6, 10 | 72.6, 8 | 75, 7.4 | 72.7, 8.5 |
| Women (%) | 59,3 | 54 | 65,1 | 57,7 |
| MMSE (Mean, SD) | 29.4, 1.1 | 25.7, 3.2 | 20.7, 4.5 | 24.4, 4.4 |
| Years education (Mean, SD) | 14, 1.1 | 8.3, 3.2 | 6.8, 4.5 | 8.2, 4.4 |
| BMI (Mean, SD) | 26.5, 3.9 | 27, 4.2 | 26.9, 4.8 | 26.9, 4.4 |
| **APOE genotype** |  |  |  |  |
| ε2ε2 (N, %) | 1 (1.1%) | 2 (0.3%) | 1 (0.3%) | 4 (0.3%) |
| ε2ε3 (N, %) | 14 (15.4%) | 50 (6.3%) | 26 (6.8%) | 90 (7.1%) |
| ε2ε4 (N, %) | 3 (3.3%) | 13 (1.6%) | 7 (1.8%) | 23 (1.8%) |
| ε3ε3 (N, %) | 56 (61.5%) | 469 (59.1%) | 210 (55.1%) | 735 (58.1%) |
| ε3ε4 (N, %) | 14 (15.4%) | 219 (27.6%) | 119 (31.2%) | 352 (27.8%) |
| ε4ε4 (N, %) | 3 (3.3%) | 41 (5.2%) | 18 (4.7%) | 62 (4.9%) |
| **CSF AD biomarkers** |  |  |  |  |
| p-tau181 (Mean, SD) | 44, 1.1 | 72.6, 3.2 | 93.7, 4.5 | 76.9, 4.4 |
| Aβ42 (Mean, SD) | 1089.2, 1.1 | 853.6, 3.2 | 695.6, 4.5 | 823.1, 4.4 |
| A-T- (N, %) | 68 (74.7%) | 251 (31.6%) | 69 (18.2%) | 388 (30.7%) |
| A-T+ (N, %) | 7 (7.7%) | 141 (17.8%) | 52 (13.7%) | 200 (15.8%) |
| A+T- (N, %) | 7 (7.7%) | 119 (15.0%) | 66 (17.4%) | 192 (15.2%) |
| A+T+ (N, %) | 9 (9.9%) | 283 (35.6%) | 193 (50.8%) | 485 (38.3%) |
| N/A (N, %) |  |  | 1 | 1 |

### 3.2 DNA Sequencing of the *APOE* locus in the Spanish Population

We identified 50 SNVs across the 3939 bp amplified *APOE* region, spanning from the promoter to the 3’UTR (Supp. Table 5). Comparison of variant calls derived from NGMLR– and GraphMap-aligned reads revealed three discrepancies. Two SNVs (19:44905756:G>A and 19:44906340:G>T) were detected exclusively in Graphmap and NGLMR alignments, respectively, across 21 and 3 chromosomes. Both variants were located within homopolymer regions and were absent from dbSNP, strongly suggesting they represented sequencing artifacts. These calls were removed from further analysis. The third discrepancy involved rs147236548 detected in a single chromosome in Graphmap-aligned reads only. This discordancy was resolved using TOPMed-imputed genome-wide array data from the GR@ACE cohort^35–37^, which supported the Graphmap call in this individual sample. Accordingly, Graphmap-aligned calls were retained, resulting in a final set of 48 SNVs for downstream analysis (Table 2).

**Table 2.**
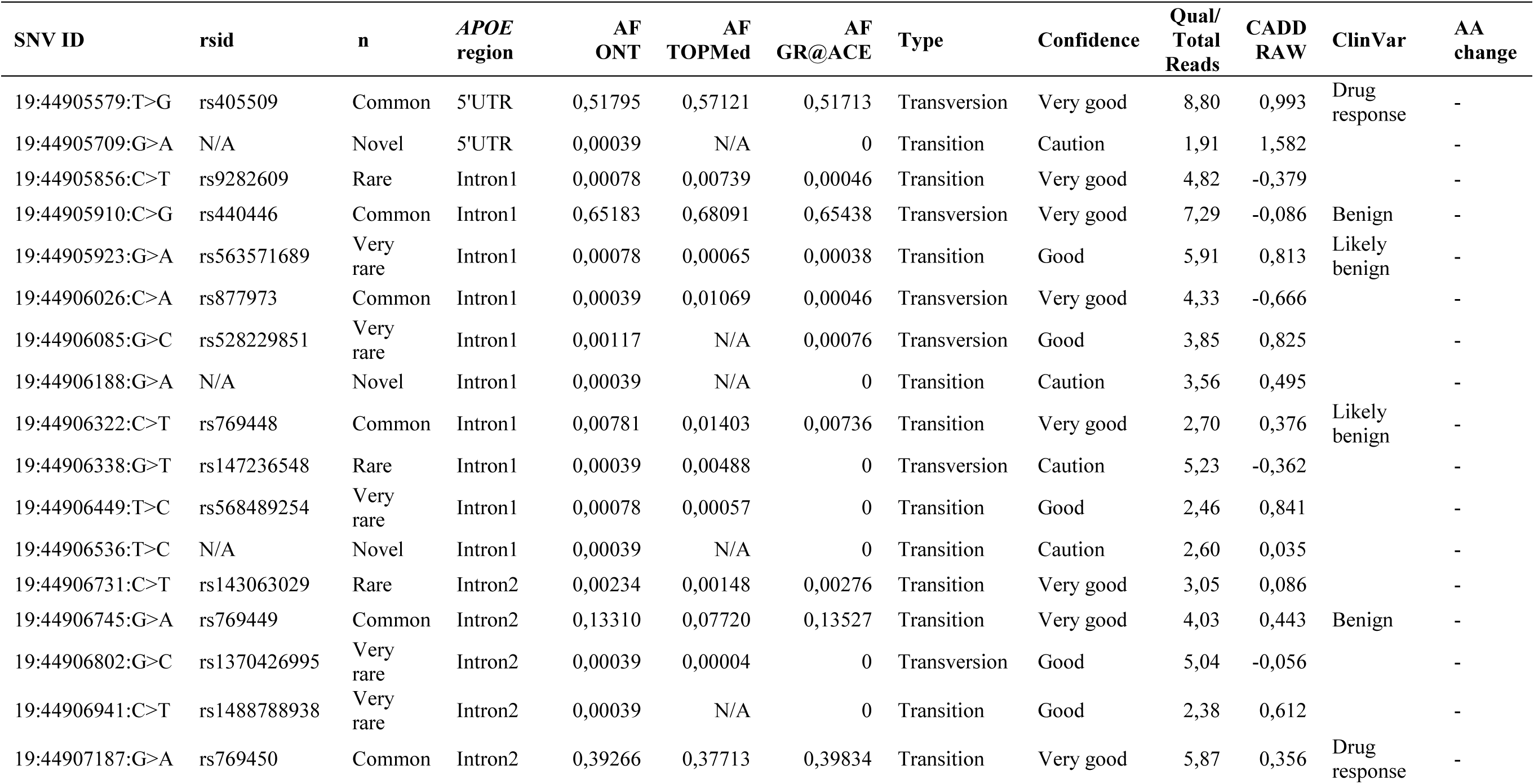
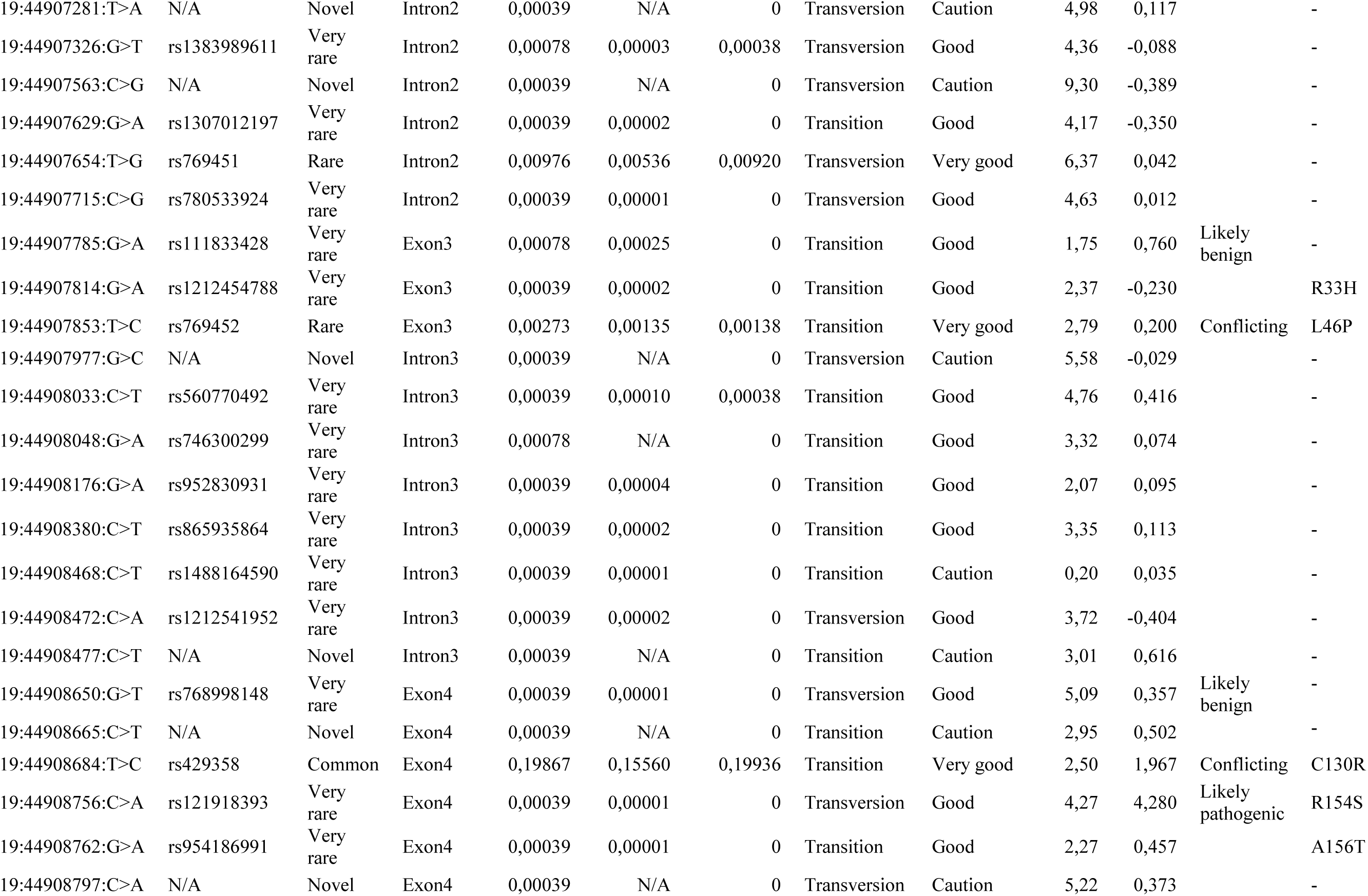
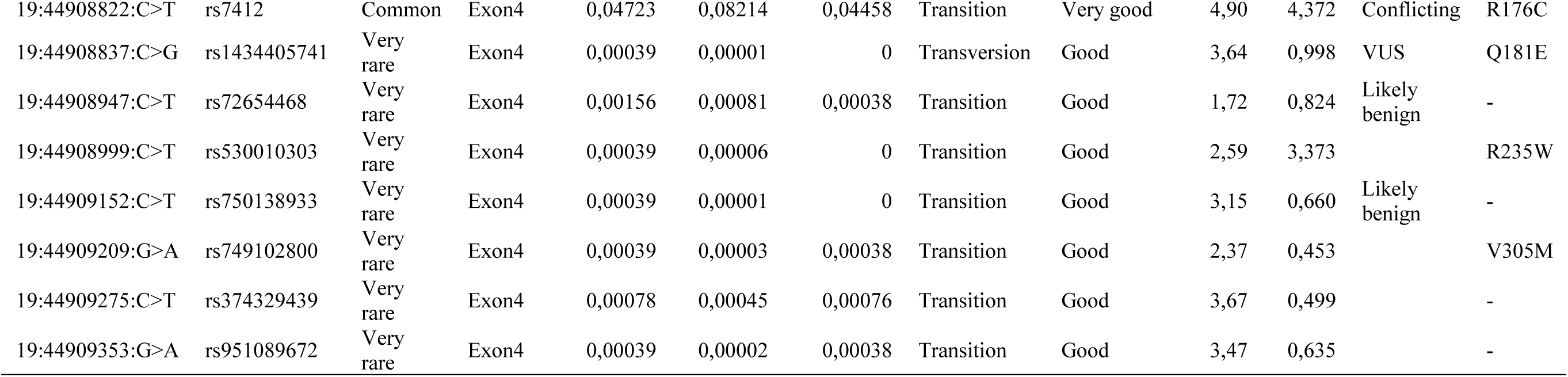
Summary of single nucleotide variants observed in the *APOE* gene region using ONT sequencing. rsIDs, genomic coordinates, reference alleles and alternative alleles are based on dbSNP build151 (GRCh38.p7). The *APOE* region is based on chromosomal coordinates of the canonical *APOE* MANE_Select transcript (ENST00000252486.9). ClinVar categories were collapsed (see methods). Aminoacid substitutions were predicted by VEP. AF ONT, alternative allele frequency in the ONT experiment; AF TOPmed, TOPMed alternative allele frequency (as reported by dbSNP build 151); AF GR@ACE, alternative allele frequency in 1026 overlapping individuals from the GR@ACE^35–37^ cohort; Qual/TotalReads, Mean QUAL to TotalReads ratio.

We classified SNVs according to their frequency in the TOPMed^52^ population and novelty status (Fig. 1, Table 2): 8 were common (MAF>0.01), 7 rare (0.01>MAF>0.001), 24 very rare (MAF<0.001), and 9 were novel variants (unreported in public databases dbSNP^53^ and gnomAD^54^) possibly representing newly identified mutations. Overall, 31 SNVs were transitions and 17 were transversions (Ts/Tv= 1.82). We further assigned confidence ratings based on allele frequency and sequencing quality metrics (Supp. Table 6). 37 variants were classified as ‘Good’ or ‘Very good’, whereas all novel SNVs were conservatively labeled as ‘Caution’ (Fig. 1, Table 2). Independent validation (e.g. Sanger sequencing) of novel variants would help rule out amplification or sequencing artifacts, consolidating these SNVs in the Spanish population. Two additional variants were labeled as ‘Caution’: rs1488164590 due to its occurrence in a homopolymer region, extremely low QUAL/Total_reads ratio, and presence in a single chromosome; and rs147236548, due to its detection by only one aligner (Table 2).

**Figure 1.**
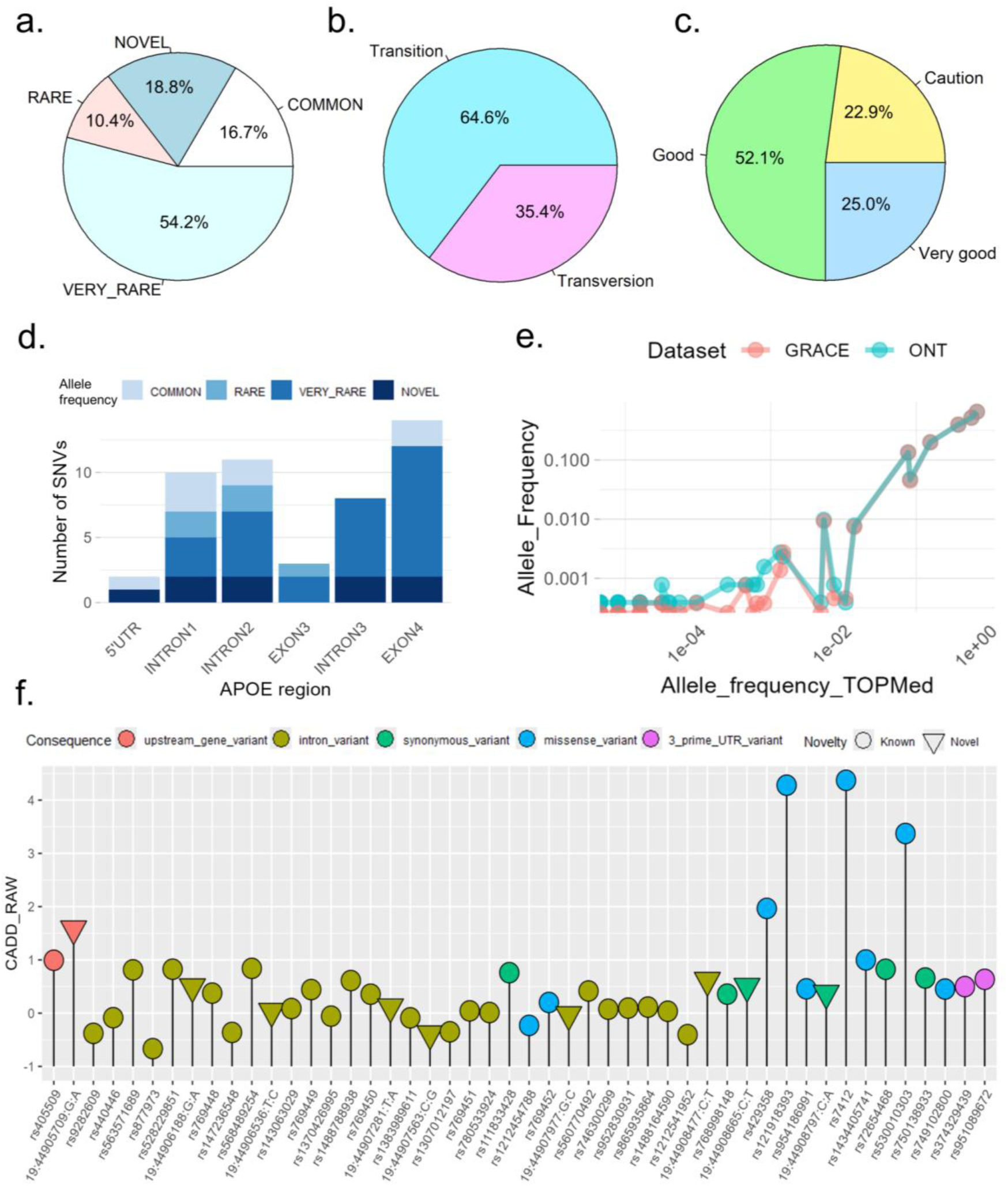
Summary of variants identified in the *APOE* locus using ONT sequencing, summarizing: **a.** variant class based on frequency and novelty status based on dbSNP annotations; **b.** substitution types; **c.** confidence annotation and; **d.** location respective to the APOE gene. **e.** Comparison of common variant frequencies Across ONT Results, GR@ACE samples and the TOPMed database. The GR@ACE data reflects the frequencies in 1026 overlapping individuals genotyped using the Affymetrix Axiom Spain Biobank array and imputed with the TOPMed reference panel, as described elsewhere^35–37^. TOPMed frequencies reflect those reported by dbSNP build 151. **f.** CADD scores for each observed variant in the *APOE* gene. Variants were sorted by genomic position and the scores are plotted alongside their corresponding consequence annotations predicted by VEP.

The observed and expected frequencies of common variants (MAF > 0.01) showed near-perfect concordance between ONT-based calls and array data from the GR@ACE cohort (Fig. 1e, Table 2), supporting the reliability of SNVs detection using our approach. In contrast, rare variants were observed at higher frequencies in ONT data compared to TOPMed-imputed genotypes, highlighting the increased sensitivity of sequencing for detecting low-frequency variation, likely reflecting incomplete haplotype representation in reference panels. The majority of very rare variants (MAF < 0.001) were singletons, consistent with their low representation in global datasets and suggesting potential population-specific variation. Notably, aside from the two canonical SNVs defining APOE ε4 and ε2 alleles (rs429358 and rs7412), all variants identified within intron 3 and exon 4 were either very rare or novel, suggesting strong sequence conservation in these regions.

Among the 9 novel variants, 6 were located within intronic regions, 1 in the 5’UTR region, and 2 are in exon 4 (Table 2). Both exonic variants (19:44908665:C>T and 19:44908797:C>A) were synonymous and therefore not expected to alter the amino acid sequence, although potential effects on splicing or regulatory elements cannot be excluded.

### 3.3 Functional annotation of identified *APOE* variants

We annotated the 48 identified variants using the Ensembl Variant Effect Predictor (VEP) tool^40^ (Supp. Table 7). Nine variants were predicted to encode missense mutations, leading to aminoacid substitutions in the APOE protein sequence. Two of these were common variants (rs429358 and rs7412), while the other seven were rare or very rare. We leveraged CADD scores to predict variant deleteriousness (Fig. 1f), highlighting three variants with the highest CADD values in the region: rs121918393 (p.Arg154Ser; APOE Christchurch), rs7412 (p.Arg176Cys), and rs530010303 (p.Arg235Trp). However, all observed scores were modest (range: 5–10), falling below commonly used thresholds for moderate or high deleteriousness (CADD ≥15–20)^55^, suggesting limited predicted functional impact based solely on in-silico metrics. Importantly, novel variants had low predicted deleteriousness.

Moreover, the SNV corresponding to the Christchurch mutation (rs121918393) was identified as as likely pathogenic based on ClinVar^56^ annotations. This variant affects an evolutionarily conserved residue within the receptor-binding domain of APOE, significantly diminishing APOE receptor binding capacity^57^, and is associated with familial hyperlipidemia^58^. Homozygosity in this variant has been linked to delayed onset of cognitive impairment in a *PSEN1* mutation carrier⁵⁸, suggesting a potential protective effect against AD, whereas heterozygosity does not appear to confer protection.

### 3.4 Long-read haplotype phasing of *APOE* SNVs

Long-read phasing using WhatsHap^41^ identified 59 distinct *APOE* haplotypes, with frequencies ranging from 38.1% (haplotype 1; n=970) to singletons (0.04%; Fig. 2; Supp. Table 8). A total of 17 haplotypes were observed three or more times, accounting for 92.74% of all chromosomes. Importantly, our results revealed that both ε3 and ε4 alleles segregate into two common haplotypes defined by the promoter variant rs405509 (Fig. 2). The two most frequent haplotypes corresponded to ε3 (ε3a and ε3b; haplotypes 1 and 2), followed by ε4 (ε4a and ε4b; haplotypes 3 and 4), whereas a single common haplotype was observed for ε2 (haplotype 5). To simplify downstream analyses, rarer haplotypes were grouped into A and B haplogroups based on their phase with rs405509 relative to the canonical *APOE* alleles (Fig. 2, Supp. Table 8).

**Figure 2:**
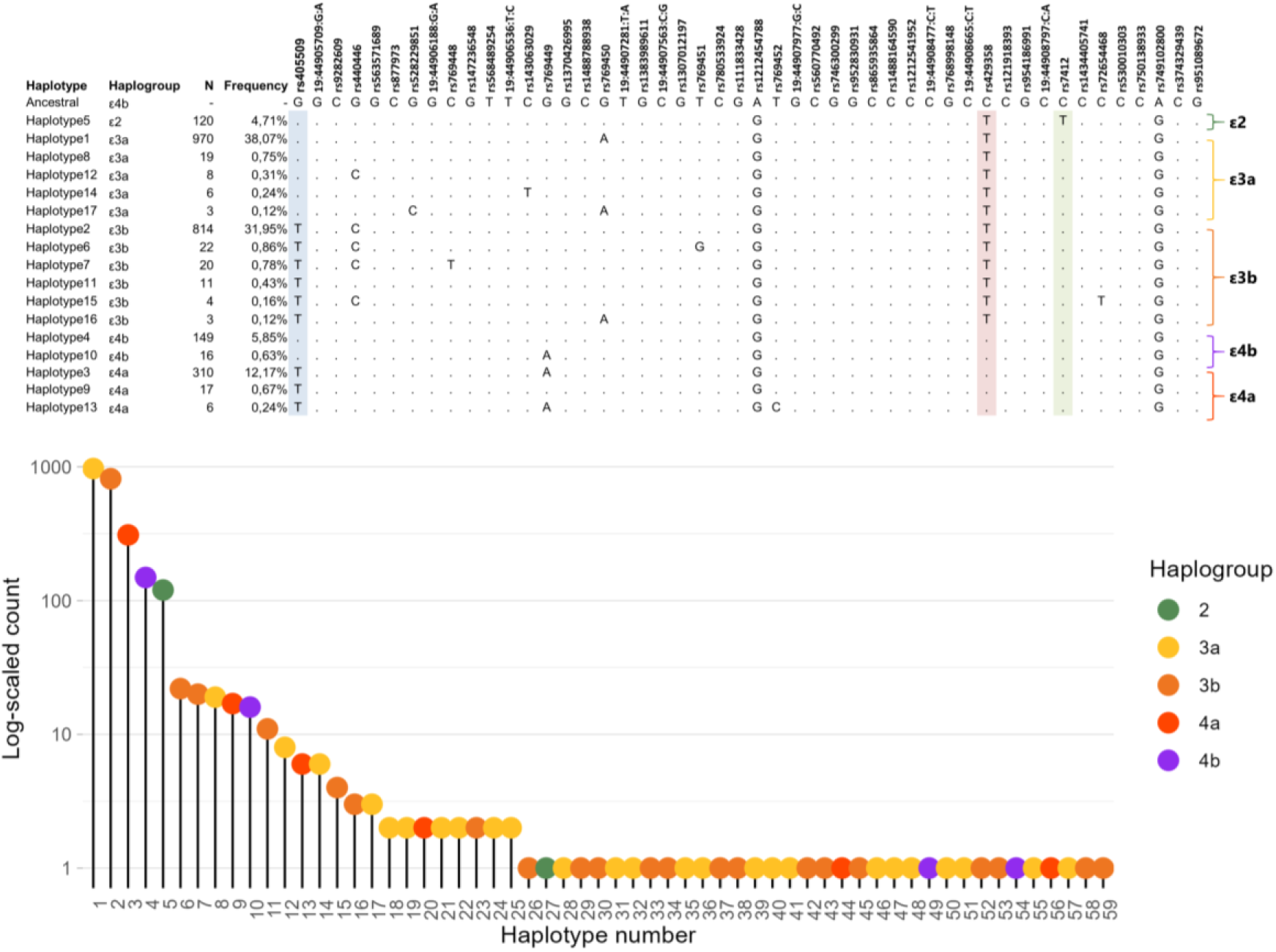
*APOE* haplotype structure in the Spanish Population. **a.** Alignment of the top 17 haplotypes (those with 3 or more occurrences) in our study population (N=1,266) with respect to the ancestral human sequence according to EMBL. Our analysis reveals five major haplogroups (ε2, ε3a, ε3b, ε4a and ε4b), determined by the phasing of the homoplasic SNV rs405509, rs429358 (the SNV determining the APOE ε4 isoform) and rs7412 (the SNV determining the APOE ε2 isoform). Haplogroups were sorted based on the presence/absence of an alternative allele in SNV rs405509, rs429358 and rs7412. Haplotypes were further sorted by their relative frequency within their haplogroup context. **b.** Frequency of the 59 identified *APOE* haplotypes displayed as the log-scaled count.

The predominance of a limited number of common haplotypes alongside a long tail of rare configurations indicates a structured haplotype architecture at the *APOE* locus. The consistent partitioning of ε3 and ε4 alleles into two major haplotypic backgrounds underscores the contribution of promoter variation to *APOE* genetic diversity. Notably, a similar haplotypic structure was previously reported in a smaller cohort using allele-specific amplification¹⁹, supporting the robustness of this pattern. This recurrent configuration is consistent with historical recombination or gene conversion events involving promoter variants, although this mechanism cannot be directly inferred from the present data.

### 3.5 Phylogenetic analysis of *APOE* haplotype sequences

To investigate the evolutionary relationships between *APOE* haplotypes, we performed a maximum likelihood phylogenetic analysis using the Tamura–Nei model implemented in *MEGA*^43^. The analysis included 59 observed haplotypes and the inferred Homo sapiens ancestral *APOE* sequence obtained from Ensembl^45^. The resulting tree (log likelihood = −404.80) was scaled by genetic distance, with branch lengths representing substitutions per site (Fig. 3).

**Figure 3:**
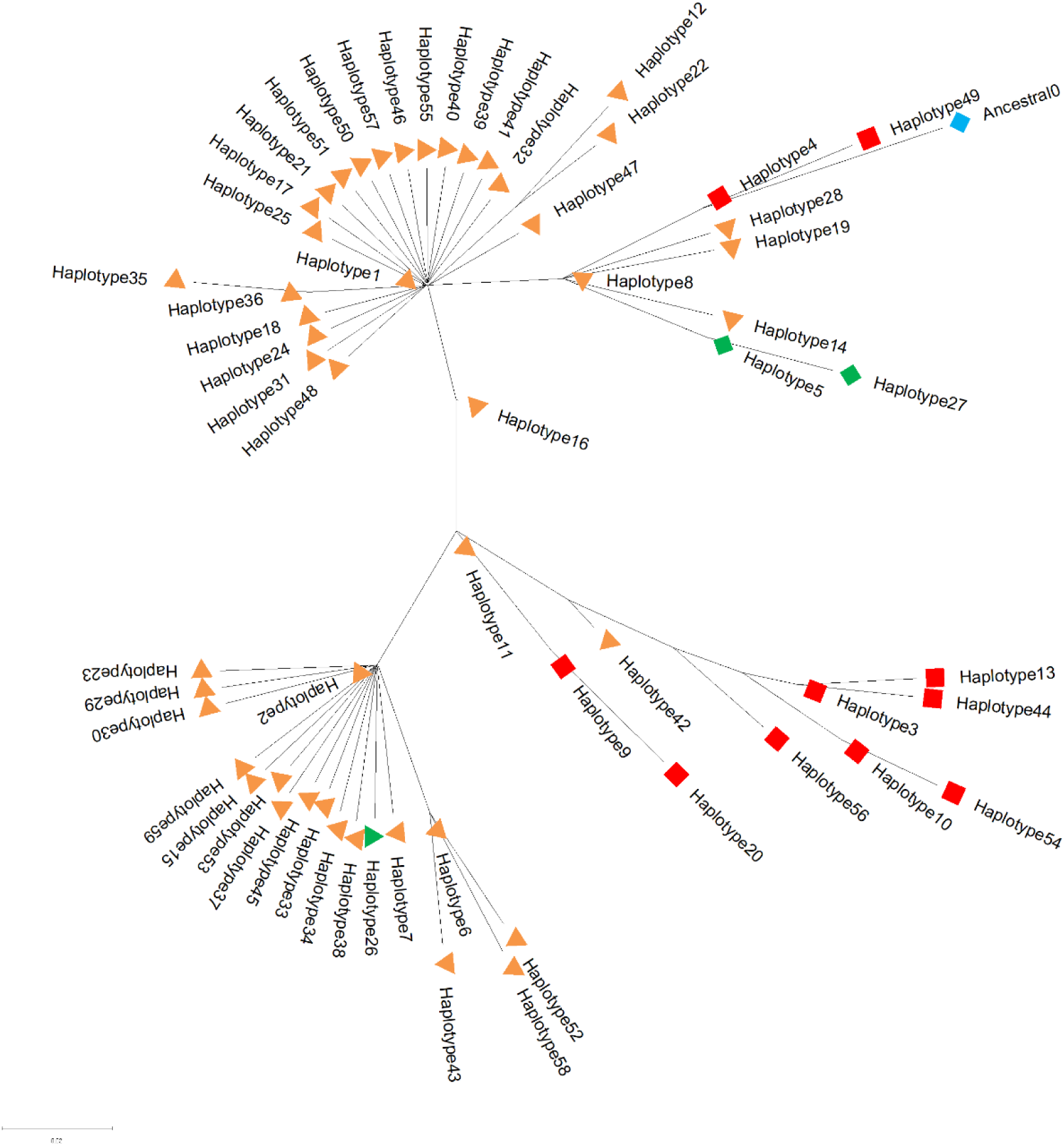
Phylogenetic Network of *APOE* intragenic Haplotypes Based on SNV Variations in the Spanish Population. This phylogenetic network illustrates the evolutionary relationships among the 59 *APOE* haplotypes identified in the Spanish population. Each node represents a distinct haplotype, and branches reflect the genetic distances based on single nucleotide variant (SNV) differences. The shapes and colors of the nodes correspond to specific isoform backgrounds: Orange triangles: ε3 background. Green triangle: Christchurch mutation under the isoform ε3 background. Green diamond: Haplogroup 2 (ε2 background). Red squares: ε4 background). Blue diamond is the ancestral allele sequence according to EMBL database. The clustering of nodes indicates shared evolutionary paths, highlighting how specific haplotypes are grouped based on shared genetic variations.

The inferred topology revealed a structured clustering of haplotypes consistent with the previously defined haplogroups. Notably, haplotype 4 (ε4b) occupied the most basal position among haplotypes observed in the Spanish population (Fig. 3), in agreement with prior reports in smaller cohorts^19^. The phylogenetic tree displays how diversification of haplotypes occurred within each observed haplogroup, resulting in variants that differ by only 1-3 positions from the most common haplotypes (Haplotypes 1-5). The phylogenetic framework also enabled assignment of rare coding variants to specific haplotypic backgrounds. For example, the *APOE* Christchurch variant (rs121918393)^59–61^ was observed on haplotype 26 within the ε3b background (Supp. Table 7), illustrating how rare functional variants are embedded within defined haplotype structures.

An intriguing aspect of our findings was the absence of a full ancestral sequence among the Spanish *APOE* haplotypes examined (Fig. 2, Fig. 3). Instead, our analysis indicated that all haplotypes, except for two singletons, harbored alternative sequences shared to Neanderthal-derived sequences at two specific positions within the gene (rs1212454788 and rs749102800, Supp. Table 8). This observation raises the possibility of introgression, suggesting that some of the genetic material associated with the *APOE* sequences in modern humans may have originated from interbreeding events between early humans and Neanderthals. However, extended sequencing is needed to elucidate this possibility.

### 3.6 Exploring *APOE* haplogroups effects on AD

To investigate whether *APOE* genetic effects extend beyond canonical ε alleles, we evaluated the impact of the identified common haplogroups (ε2, ε3a, ε3b, ε4a, ε4b) on disease progression and core AD biomarkers. Given the structured haplotype architecture at this locus, we hypothesized that haplotypic background may modulate APOE-associated risk.

#### 3.6.1 Effect on disease progression

We assessed the effect of *APOE* haplogroups on progression from MCI to AD dementia in 724 individuals with baseline MCI and longitudinal follow-up (Supp. Table 4). Individuals were initially stratified into haplogenotypes based on combinations of the five common haplogroups (ε2, ε3a, ε3b, ε4a, ε4b). An overview of Kaplan-Meier (KM) curves revealed the expected dose-dependent effect of ε4 on conversion rates and the protective effect of ε2 (Fig. 4a). Importantly, differences in survival curves attributable to haplogenotype dosage were also observed within subjects with the same *APOE* genotype (Fig. 4b–c). Log-rank tests revealed a borderline significant association between the ε3b haplotype and slower disease progression in ε3ε3 subjects (*p=*0.057), and a significant association of the ε4a haplotype with slower progression in ε3ε4 (*p=*0.020) and ε4ε4 (*p=*0.028) individuals. Together, these patterns suggested a modest protective effect of ε3b within ε3 carriers and a stronger protective effect of ε4a among ε4 carriers.

**Figure 4.**
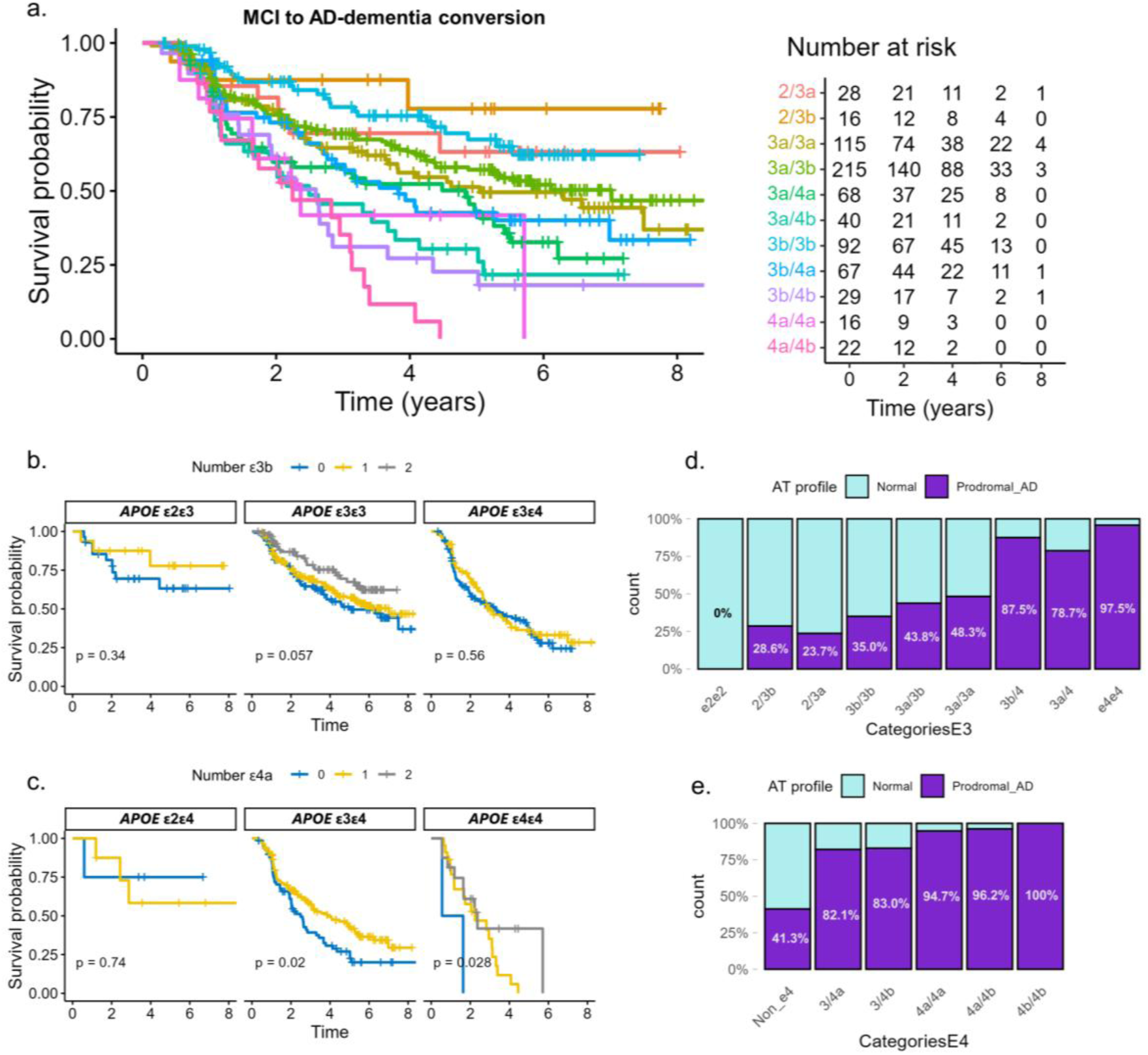
Effects of *APOE* haplogroups on AD progression and core biomarkers. **a.** KM plot displaying survival curves of MCI to AD-dementia progression for subjects with different combinations of *APOE* haplogenotypes. Analysis was restricted to haplogenotype categories with 25 or more occurrences. The p-values displayed are derived from log-rank tests comparing the survival curves between groups. The number of non-converted, uncensored individuals after 2, 4, 6 and 8-year follow-up is also shown. **b-c.** Stratified KM plots showing the effect of the e3b and e4a haplogenotypes in disease progression, respectively. **d-e.** Proportion of A+T+ versus A-T-subjects with different e3 and e4 haplogenotype combinations, respectively.

To formally test these observations, we performed Cox proportional hazards models adjusted by sex, age and baseline MMSE under three complementary frameworks: (i) a joint model including *APOE* ε2 and ε4 allele dosage together with ε3b and ε4a haplogenotype dosage; (ii) genotype-stratified analyses followed by inverse-variance meta-analysis; and (iii) interaction models testing modification of ε4 effects by ε3b and ε4a (see methods and Supp. Table 3). Across analyses, ε4a haplogenotype dosage was consistently associated with reduced risk of progression compared to ε4b (Table 3), reaching statistical significance in the joint model (*HR[95%CI]=*0.63[0.46–0.88], *p=*0.007, *N=*700), as well as in the ε3ε4 (*HR[95%CI]=*0.60[0.41–0.88], *p*=0.008, *N=*197) and ε4ε4 strata (*HR[95%CI]=*0.39[0.16–0.90], *p=*0.028, *N=*39), and in the interaction analysis (*HR[95%CI]=*0.60[0.41–0.87], *p=*0.007, *N=*606). In contrast, although ε3b dosage showed effect directions consistent with a protective effect, none of the analyses reached statistical significance (Table 3).

**Table 3.**
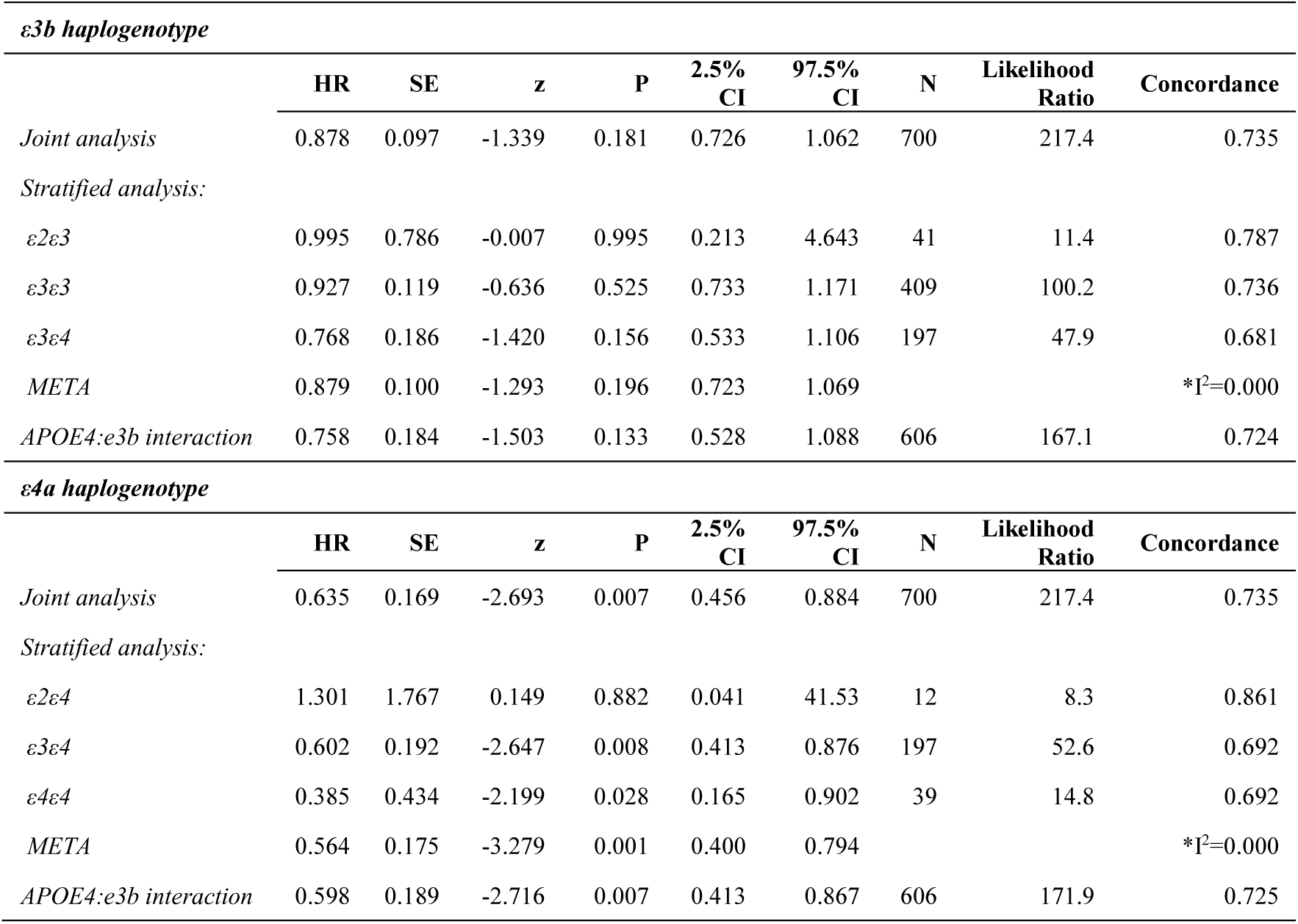
Cox proportional hazards model results for MCI to AD dementia progression. Models were adjusted by age at baseline, sex and baseline MMSE.

| *ε3b haplogenotype* |  |  |  |  |  |  |  |  |  |
| --- | --- | --- | --- | --- | --- | --- | --- | --- | --- |
|  | HR | SE | z | P | 2.5%<br>CI | 97.5%<br>CI | N | Likelihood<br>Ratio | Concordance |
| *Joint analysis* | 0.878 | 0.097 | -1.339 | 0.181 | 0.726 | 1.062 | 700 | 217.4 | 0.735 |
| *Stratified analysis:* |  |  |  |  |  |  |  |  |  |
| *ε2ε3* | 0.995 | 0.786 | -0.007 | 0.995 | 0.213 | 4.643 | 41 | 11.4 | 0.787 |
| *ε3ε3* | 0.927 | 0.119 | -0.636 | 0.525 | 0.733 | 1.171 | 409 | 100.2 | 0.736 |
| *ε3ε4* | 0.768 | 0.186 | -1.420 | 0.156 | 0.533 | 1.106 | 197 | 47.9 | 0.681 |
| *META* | 0.879 | 0.100 | -1.293 | 0.196 | 0.723 | 1.069 |  |  | *I<sup>2</sup>=0.000 |
| *APOE4:ε3b interaction* | 0.758 | 0.184 | -1.503 | 0.133 | 0.528 | 1.088 | 606 | 167.1 | 0.724 |
| *ε4a haplogenotype* |  |  |  |  |  |  |  |  |  |
|  | HR | SE | z | P | 2.5%<br>CI | 97.5%<br>CI | N | Likelihood<br>Ratio | Concordance |
| *Joint analysis* | 0.635 | 0.169 | -2.693 | 0.007 | 0.456 | 0.884 | 700 | 217.4 | 0.735 |
| *Stratified analysis:* |  |  |  |  |  |  |  |  |  |
| *ε2ε4* | 1.301 | 1.767 | 0.149 | 0.882 | 0.041 | 41.53 | 12 | 8.3 | 0.861 |
| *ε3ε4* | 0.602 | 0.192 | -2.647 | 0.008 | 0.413 | 0.876 | 197 | 52.6 | 0.692 |
| *ε4ε4* | 0.385 | 0.434 | -2.199 | 0.028 | 0.165 | 0.902 | 39 | 14.8 | 0.692 |
| *META* | 0.564 | 0.175 | -3.279 | 0.001 | 0.400 | 0.794 |  |  | *I<sup>2</sup>=0.000 |
| *APOE4:ε3b interaction* | 0.598 | 0.189 | -2.716 | 0.007 | 0.413 | 0.867 | 606 | 171.9 | 0.725 |

#### 3.6.2 Effect on core AD biomarkers

We next applied this framework to explore whether the *APOE* haplotypes were associated with AD pathology as measured by CSF Aβ42 and p-tau181 levels. To test our hypothesis, we ran logistic models comparing subjects with a A+T+ profile with those with a A-T-profile, adjusted by age and sex (Supp. Table 9). Joint model results indicated that *APOE*, age and sex explained a large fraction of the variance in AT classification (Nagelkerke pseudo-R^2^ = 0.413). However, no significant associations were observed between ε4 haplogenotypes and AD biomarker positivity. In contrast, the ε3b haplogenotype showed a borderline significant protective effect (*OR[95%CI]=*0.76[0.57–1.00], *p=*0.053, *N=*679). This effect was not observed in ε3ε4 individuals, suggesting that the presence of an ε4 allele substantially overrides the potential protective effect exerted by the ε3b subhaplotype.

### 3.7 Effect of *APOE* haplogroups in CSF APOE protein levels

To investigate potential mechanisms underlying haplotype-specific effects, we examined the effect of the *APOE* haplogroups on CSF APOE protein levels. Given that the genetic variation differentiating ε3 and ε4 haplogroups is located at the promoter region, we hypothezised that haplotypic background may modulate APOE expression in the central nervous system in an allele-dependent manner.

Although Somascan APOE measurements were available for a large proportion of the samples, previous work has shown that these measures have very low reproducibility and are thus unreliable^62^. Therefore, we retrieved Olink CSF APOE measurements available for 480 samples^46^, and complemented these with Lumipulse-based quantification in additional samples with available CSF.

*APOE* genotype alone explained a substantial proportion of the variance in CSF APOE levels in in both Olink (adjusted R^2^ = 0.47) and Lumipulse (adjusted R^2^ = 0,48) experiments (Supp Fig. 2-3, Supp. Table 10), replicating the well-stablished gradient of decreasing APOE levels across genotypes (ɛ2 > ɛ3 > ɛ4). Inclusion of covariates capturing inter-individual variability in CSF turnover (OPCML levels) and blood-brain barrier integrity (second proteomic principal component)^50^ markedly improved model prediction in Olink (adjusted R^2^ = 0.83) and Lumipulse data (adjusted R^2^ = 0.85), explaining over 80% of the variance in APOE levels, and improving the signal-to-noise ratio (Supp Fig. 2–3). Notably, inclusion of age and sex as covariates did not further improve model performance (Supp. Table 10).

Pairwise comparisons between haplogenotypes within the same *APOE* allelic context revealed no significant differences in CSF APOE levels, indicating that haplotypic background has minimal impact on overall APOE protein abundance beyond the effect of ε alleles (Fig. 5). Consistently, regression analyses using the same modeling framework as for disease progression showed no robust associations between haplogroups and APOE levels (Supp. Table 11). A nominal association was observed in Olink data for ε3b in ε3ε4 individuals (*β[95%CI]=*0.0996[0.0085–0.1907], *p=*0.032, *N=*125) and in interaction models (*β[95%CI]=*0.0994[0.0136–0.1851], *p=*0.023, *N=*410), suggesting a potential increase in APOE levels in the presence of an ε4 allele; however, this effects were not replicated in the Lumipulse dataset (Supp. Table 11). Furthermore, analysis of isoform-specific APOE4 levels (available only in Lumipulse data) showed no evidence of modulation by ε4 haplogroups either (Fig. 5). Overall, these results indicate that promoter-driven haplotypic variation at the *APOE* locus does not substantially influence CSF APOE protein levels, and therefore this endophenotypes does not account for the haplotype-dependent effects observed on disease progression.

**Figure 5.**
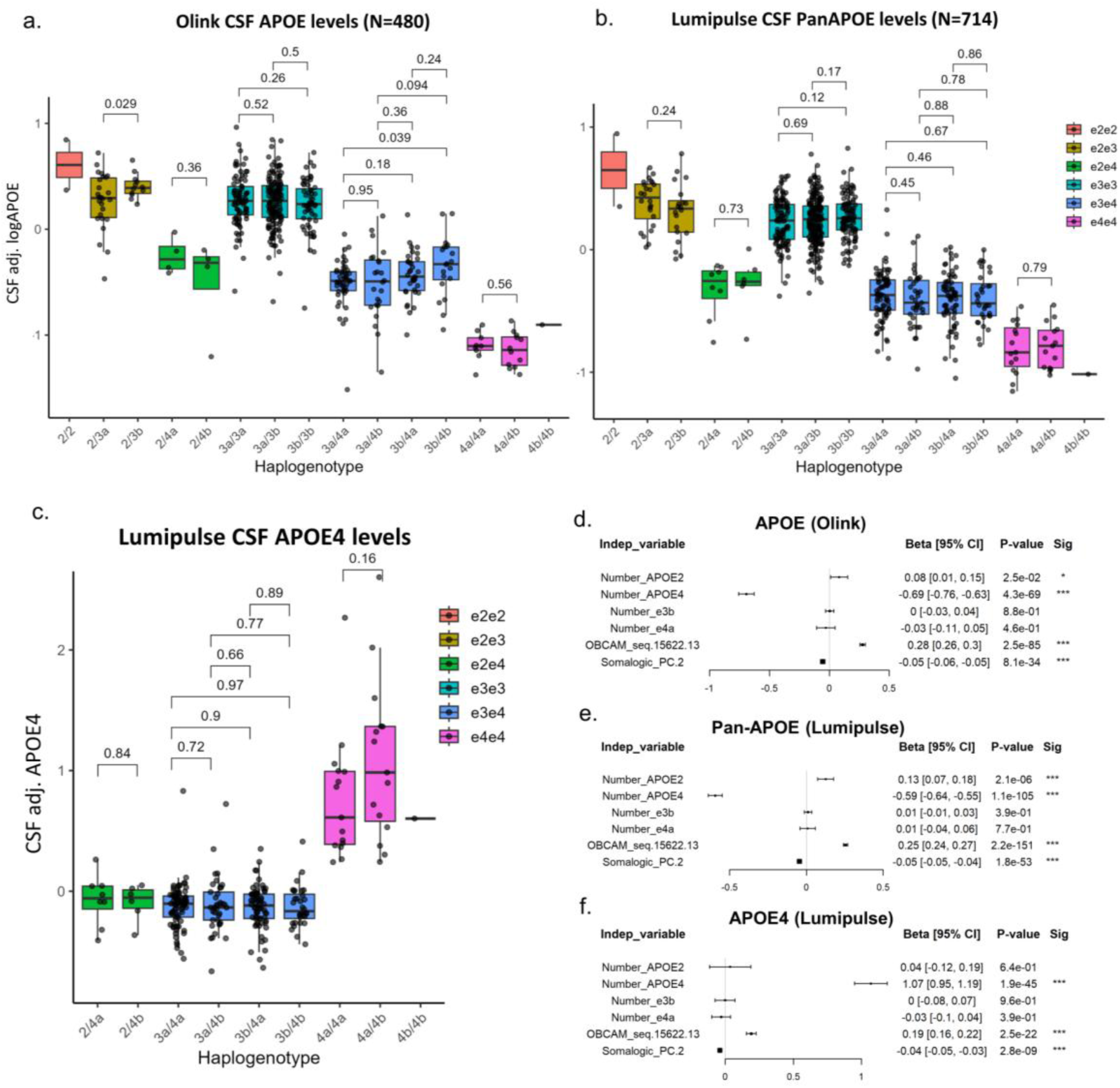
Effect of APOE alleles and haplogenotypes on CSF APOE protein levels. **a-c.** CSF levels of Olink-based APOE (**a**), Lumipulse-based Pan-APOE (**b**), and Lumipulse-based APOE4 isoform (**c**) for each haplogenotype combination. Bars show p-values derived from pairwise t-test comparisons. **d-f.** Multivariate linear model results including the number of APOE ε4 and ε2 alleles, the number of ε3b and ε4a haplotypes, CSF OPCML (measured by the OBCAM_seq.15622.13 somamer) and the second principal component (as measured by Somascan) as predictors of CSF Olink-based APOE (**d**), Lumipulse-based Pan-APOE (**e**), and Lumipulse based APOE4 isoform (**f**).

## 4 Discussion

*APOE* represents the strongest genetic risk locus for sporadic AD^4,63^, yet despite decades of research the mechanisms underlying its effects are still poorly understood. In this study, we demonstrate that *APOE* ε3 and ε4 alleles are embedded within specific haplotypic configurations that further modulate disease progression beyond the effects of the classic ε3/ε4 isoforms. In particular, we identified differential effects between ε4 haplogenotypes, with ε4a consistently associated with reduced risk of progression compared to ε4b. Additionally, we report two borderline significant protective associations of the ε3b haplogenotype in ε3ε3 subjects for AD pathology, as determined by CSF core AD biomarkers, and for MCI to AD progression rates, determined by analysis of KM curves.

Our findings suggest that the combination of intragenic regulatory variants may fine-tune *APOE* expression. This is supported by our observation that the ɛ4a and ɛ3b haplogroups exert opposing effects despite sharing a common genetic background (the homoplasic rs405509-T allele located in the promoter region) in conjunction with the ε4 and ε3 alleles, respectively. This could point to a theoretical mechanism of action, by which enhanced expression of the ɛ3 isoform could be slightly protective, while enhanced expression of the pathogenic ɛ4 isoform could lead to increased risk. Notably, each haplogroup is uniquely combined with additional SNVs in intronic enhancers of the gene (Fig. 2), including the rs440446-C allele in most ɛ3B haplogroup haplotypes, the rs769449-A allele in the dominant ɛ4a haplotype (Haplotype 3), and the rs769450-A allele in Haplotype1 (ɛ3a). It is possible that the specific combination of these variants within enhancer-active regions may induce paradoxical effects on the gene expression of homoplasic variants. However, an exhaustive functional characterization of the entire intragenic haplotypes is necessary to understand the mechanisms associated with our findings.

Interestingly, despite the strong effect of *APOE* genotype in CSF APOE levels (Fig. 5), no differences attributed to haplogenotypes were identified in our study. This was unexpected, given that the homoplasic rs405509 variant is located in the promoter region, which led us to hypothesize that the observed haplotypic effects on disease progression and biomarker status might be mediated through differential *APOE* expression. Instead, our results suggest that the mechanisms underlying haplotype-dependent effects are largely independent of those driving global CSF APOE levels. It is important to note that APOE abundance in our study was quantified using affinity-based platforms (Olink, Lumipulse). As APOE isoforms (ε2, ε3 and ε4) exhibit substantial differences in lipidation status and receptor-binding efficiency^64,65^, these biochemical differences may influence quantification independently of true protein concentration. Consequently, subtler effects of cis-acting variants (such as cell-type-specific regulation of gene expression) may be obscured by isoform-related measurement biases, potentially explaining the lack of detectable haplotype-specific differences. Future studies exploring the impact of *APOE* haplotypes on isoform expression on a single-cell level might help elucidate this question.

The existence of cis-regulatory modulators of *APOE* has been previously suggested, particularly in studies reporting ancestry-dependent variation in ε4-associated risk^6,8–10^. Several variants in the promoter and intronic regions, including rs405509 and rs769449, have been proposed to influence *APOE* expression or regulatory activity^66–69^. These findings are consistent with observations from regional and genome-wide studies showing that a relatively large number of independent variants across the locus can modulate APOE protein levels, even in a tissue-specific manner^70,71^. In addition, regulatory elements within the 3’ region of the gene (including the coding SNVs encoding the APOE isoforms themselves) been implicated in epigenetic modulation through CpG-dependent mechanisms^72–75^. Our long-read sequencing approach extends these observations, enabling direct phasing of regulatory variants with *APOE* coding alleles, resolving previous ambiguity and demonstrating that such regulatory variation is organized into recurrent haplotypic configurations.

Previous research reinforces the importance of phasing the potential modulators of *APOE* expression with APOE protein isoforms. However, due to the small scale of previously reported studies, little was known about the clinical impact of the detected sub-haplotypes or common haplogroups. The use of ONT for generating long-range haplotypes has proven instrumental in studying the *APOE* locus^22^ and other regions across the human genome^76^, although the existence of technical problems must not be ignored^77^. To our knowledge, this is the largest study indicating differential effects of sub haplotypes within the ε4 and ε3 isoforms. Our strategy has allowed the accurate phasing of the nearest upstream intragenic regulatory variants in the locus with the classic protein isoforms, resolving the homoplasy of key upstream exon 4 variants. The homoplasy observed for rs405509 could be attributed to a single ancestral recombination (or gene conversion) event within the *APOE* gene. This event likely reshuffled SNVs in the promoters and intronic enhancers of the *APOE* gene in the ancestral ɛ4b and possibly ɛ3b backgrounds, leading to the differentiation of the ε3 and ε4 isoforms into the two (A and B) haplogroups in a single mutation step.

Importantly, the haplotypic architecture identified using our method is somewhat consistent with other small-scale studies previously published. *Fullerton et al.* used allele-specific amplification techniques to resolve homoplasy in the *APOE* locus and detected ε3 and ε4 sub haplotypes across diverse populations^19^. Our findings confirm these early observations, particularly for the common haplogroups detected. More recently, *Abondio et al.* demonstrated the presence of ε3 and ε4 sub haplotypes across geographic macro areas^78^. Although their study used inference methods rather than long-read sequencing, the six core variants they identified (rs440446, rs565782572, rs769449, rs769450, rs429358, and rs7412) represent a shared haplotype cluster that aligns well with our findings. This study reinforces the relevance of long-read sequencing for characterizing *APOE* haplogroups, validating the core haplotype composition in different human populations. The study of intragenic haplogroups may be particularly relevant because they are not dependent on ancestry background. This implies that detected haplogroups are present across different ancestries, albeit in varying proportions. Furthermore, the combination of these intragenic haplogroups with other distal and more ancestry-dependent elements could complicate the interpretation of studies aimed at identifying ancestry-specific modulators of the ε4 allele. In any case, large scale studies are necessary to elucidate the intragenic haplogroup composition in different populations and its impact in ε4 expression.

From an evolutionary perspective, our findings raise intriguing questions about the evolutionary trajectory of the human *APOE* locus. Previous studies have suggested that the ε4 allele may have been positively selected in ancestral human populations due to its advantageous effects on lipid metabolism and immune response^79^. However, the emergence of sub haplotypes such as ɛ4a, which appears to mitigate some of the adverse effects of the ε4 allele, suggests a more complex evolutionary history involving balancing selection and regulatory adaptations. In addition, we identified two nucleotide variants shared with sequences reported in multiple archaic hominin genomes (Denisovan and Neanderthal), which differ from the inferred ancestral allele (Supp. Table 7). Notably, the human ancestral allele was observed only as a singleton for both variants in our dataset, whereas the derived allele was nearly fixed across individuals. One of these variants results in a Val305Met substitution (rs749102800). This pattern is consistent with a potential contribution of archaic introgression at the *APOE* locus, possibly introducing adaptations influencing APOE function in recent human evolutionary history. However, alternative explanations such as incomplete lineage sorting cannot be excluded without dedicated population genetic analyses. Further investigation incorporating extended haplotypic context and diverse populations will be required to clarify the evolutionary origin and functional relevance of these variants.

Present results may have significant implications for the design of *APOE*-targeted gene therapies. Gene-editing strategies aimed at controlling ε4 expression, such as CRISPR-Cas9 or epigenome-based approaches^80^, should carefully consider the potential for unintended effects on neighboring regulatory elements within the less harmful ɛ4a haplogroup. Likewise, therapeutic strategies that seek to upregulate ε3 expression must account for the presence of enhancers within the ɛ3b haplogroup, which could amplify ε3 isoform levels. Again, a comprehensive understanding of *APOE* haplotype structure is thus essential for the rational design of future isoform-specific interventions.

Despite the novel insights provided by our study, several limitations must be acknowledged. First, our sample size, while robust for a single-cohort study, may not capture the full spectrum of haplotypes present across diverse populations. Ancestry-specific effects, highlighted in several studies^11,81,82^, also influence the impact of these *APOE* intragenic haplogroups on AD risk. Future studies should aim to replicate our findings in larger, multi-ethnic cohorts to assess the generalizability of these results. Second, measuring APOE protein levels in CSF remains challenging, as different platforms yield varying results depending on the assay method used (e.g., Somascan, ELISA, Olink). Therefore, while our findings are robust, they should be interpreted with caution until validated using complementary methodologies. Third, our study focused exclusively on APOE protein levels in the CSF. The effects of these intragenic haplogroups in other tissues, particularly circulating APOE measured in plasma and produced in the liver, remain to be determined. A more comprehensive analysis of the impact of these intragenic haplogroups across multiple cell types and tissues is warranted. Fourth, this study centers on the *APOE* gene itself without accounting for the broader *APOE* region, which includes other potentially AD-related genes such as *TOMM40*, *BCAM*, or *APOC1*. Future research should aim to map the full haplotype structure of the entire 19q13 region to provide a more complete understanding of its functional and genetic landscape. Fifth, due to the inherent limitations of ONT, we focused our sequencing effort solely on SNV sites. Indel variants might further contribute to intragenic haplotype diversity of the *APOE* locus. Sequencing efforts combining short-read and long-read sequencing are necessary to fully characterize the exact sequence composition of *APOE* intragenic haplogroups. This information will be essential and relevant for elucidating the functional mechanisms behind the observed effects on APOE protein isoform expressivity.

In summary, our high-resolution intragenic haplotype analysis of the *APOE* gene revealed different alleles within the ε3 and ε4 backgrounds that have distinct effects on MCI to AD progression and AD biomarker status. These findings provide new insights into the functional regulation of *APOE* in AD, suggesting that precise modulation of isoform-specific expression may be a promising therapeutic strategy. Further research is needed to explore the broader implications of these haplogroups in AD risk and pathogenesis and to refine our understanding of the regulatory mechanisms that govern *APOE* function in the brain. The prevalence of specific haplotypes, alongside insights into their evolutionary trajectories and possible Neanderthal genetic contributions, opens new research avenues aimed at elucidating the genetic elements that influence the expression of *APOE* locus. Our findings lay the groundwork for further exploration of the implications of intragenic *APOE* subhaplotypes in human populations, emphasizing the importance of continued research in *APOE* regulation and its impact in AD and other human conditions.

## Data Availability

All data produced in the present study are available upon reasonable request to the authors

## Acknowledgments

The present work has been performed as part of the Doctoral thesis of Pablo García-González at the University of Barcelona (Barcelona, Spain). We would like to thank patients and controls who participated in this project. They were processed following standard operating procedures with the appropriate approval of the Ethical and Scientific Committee. Informed consents for lumbar puncture, genetic analyses, and the anonymized use of clinical records for research were approved by the Ethics Committee of the Hospital Clinic i Provincial de Barcelona (Barcelona, Spain) in accordance with Spanish biomedical laws (Law 14/2007, of July 3, on biomedical research; Royal Decree 1716/2011, of November 18). The study also adhered to the recommendations of the Declaration of Helsinki. The Harpone project, encompassing all the research described in this manuscript, has also been approved by the Ethics Committee of the Hospital de Bellvitge (Barcelona, Spain) (Acta 04/21).

## 6 Funding

Authors acknowledge the support of the Agency for Innovation and Entrepreneurship (VLAIO) grant N° PR067/21 and Janssen for the HARPONE project and the ADAPTED project the EU/EFPIA Innovative Medicines Initiative Joint Undertaking Grant N° 115975. Also, the Spanish Ministry of Science and Innovation, Proyectos de Generación de Conocimiento grants PID2021-122473OA-I00, PID2021-123462OB-I00 and PID2019-106625RB-I00. ISCIII, Acción Estratégica en Salud integrated in the Spanish National R+D+I Plan and financed by ISCIII Subdirección General de Evaluación and the Fondo Europeo de Desarrollo Regional (FEDER “Una manera de hacer Europa”) grants PI17/01474, PI19/00335, PI22/01403 and PI22/00258. The support of CIBERNED (ISCIII) under the grants CB06/05/2004 and CB18/05/00010. The support from PREADAPT project, Joint Program for Neurodegenerative Diseases (JPND) grant N° AC19/00097, and from DESCARTES project, German Research Foundation (DFG). The support of Fundación bancaria “La Caixa”, Fundación ADEY, Fundación Echevarne and Grifols SA (GR@ACE project). ACF received support from the Instituto de Salud Carlos III (ISCIII) under the grant Sara Borrell (CD22/00125). PGG was supported by CIBERNED employment plan (CNV-304-PRF-866). IdR is supported by the ISCIII under the grant FI20/00215. AR is also supported by STAR Award. University of Texas System. Tx, United States, The South Texas ADRC. National Institute of Aging. National Institutes of Heath. USA. (P30AG066546), the Keith M. Orme and Pat Vigeon Orme Endowed Chair in Alzheimer’s and Neurodegenerative Diseases (2024-2025) and Patricia Ruth Frederick Distinguished Chair for Precision Therapeutics in Alzheimer’s and Neurodegenerative Diseases (2025-2028). Part of this study was also funded by the German Federal Ministry of Education and Research (BMBF) within the EU program JPND (Grant number: PreADAPT project 01ED2007A) and by BMBF (Grant numbers: DESCARTES project 01EK2102B and 01EK2102A).

## Notes

### Competing Interest Statement

The authors have declared no competing interest.

### Author Declarations

Informed consents for lumbar puncture, genetic analyses, and the anonymized use of clinical records for research were approved by the Ethics Committee of the Hospital Clinic i Provincial de Barcelona (Barcelona, Spain) in accordance with Spanish biomedical laws (Law 14/2007, of July 3, on biomedical research; Royal Decree 1716/2011, of November 18). The study also adhered to the recommendations of the Declaration of Helsinki. The Harpone project, encompassing all the research described in this manuscript, has also been approved by the Ethics Committee of the Hospital de Bellvitge (Barcelona, Spain) (Acta 04/21).

### Summary of Updates

This revised version of the manuscript incorporates an updated analytical framework, including the joint analysis, genotype-stratified meta-analysis, and interaction models based on APOE haplotypes. We also introduce a new analysis including APOE Lumipulse measurements to validate the effect of APOE haplogroups on CSF APOE levels. All statistical analyses were conducted on R v4.1.1. In addition, the manuscript clarity and readability has been improved, and a more appropriate title has been selected.

