## Supplementary Figures for "Long-read sequencing reveals two common *APOE* ε3 and ε4 intragenic haplotypes in the Spanish population"

### Slide 1
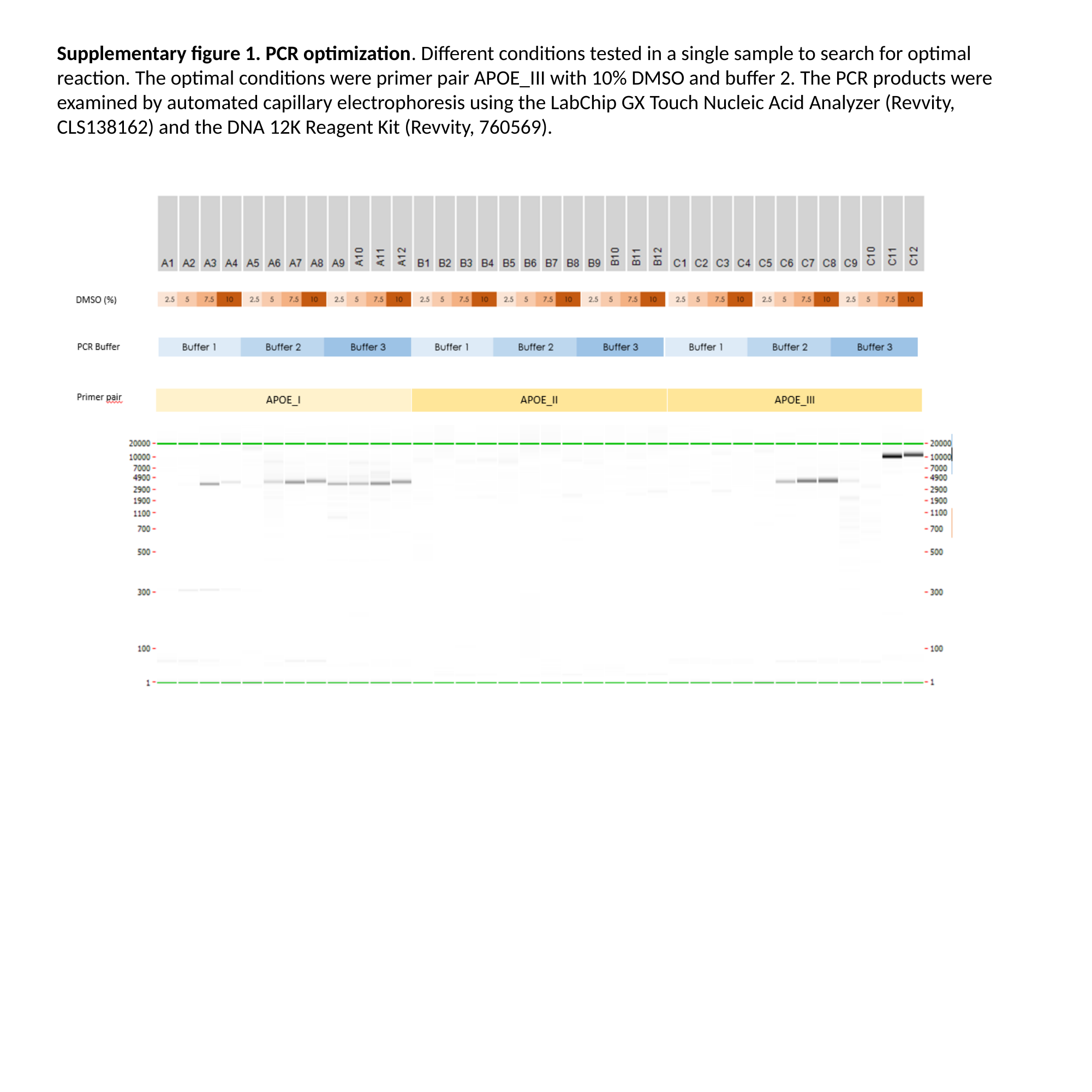

Supplementary figure 1. PCR optimization. Different conditions tested in a single sample to search for optimal reaction. The optimal conditions were primer pair APOE_III with 10% DMSO and buffer 2. The PCR products were examined by automated capillary electrophoresis using the LabChip GX Touch Nucleic Acid Analyzer (Revvity, CLS138162) and the DNA 12K Reagent Kit (Revvity, 760569).

### Slide 2
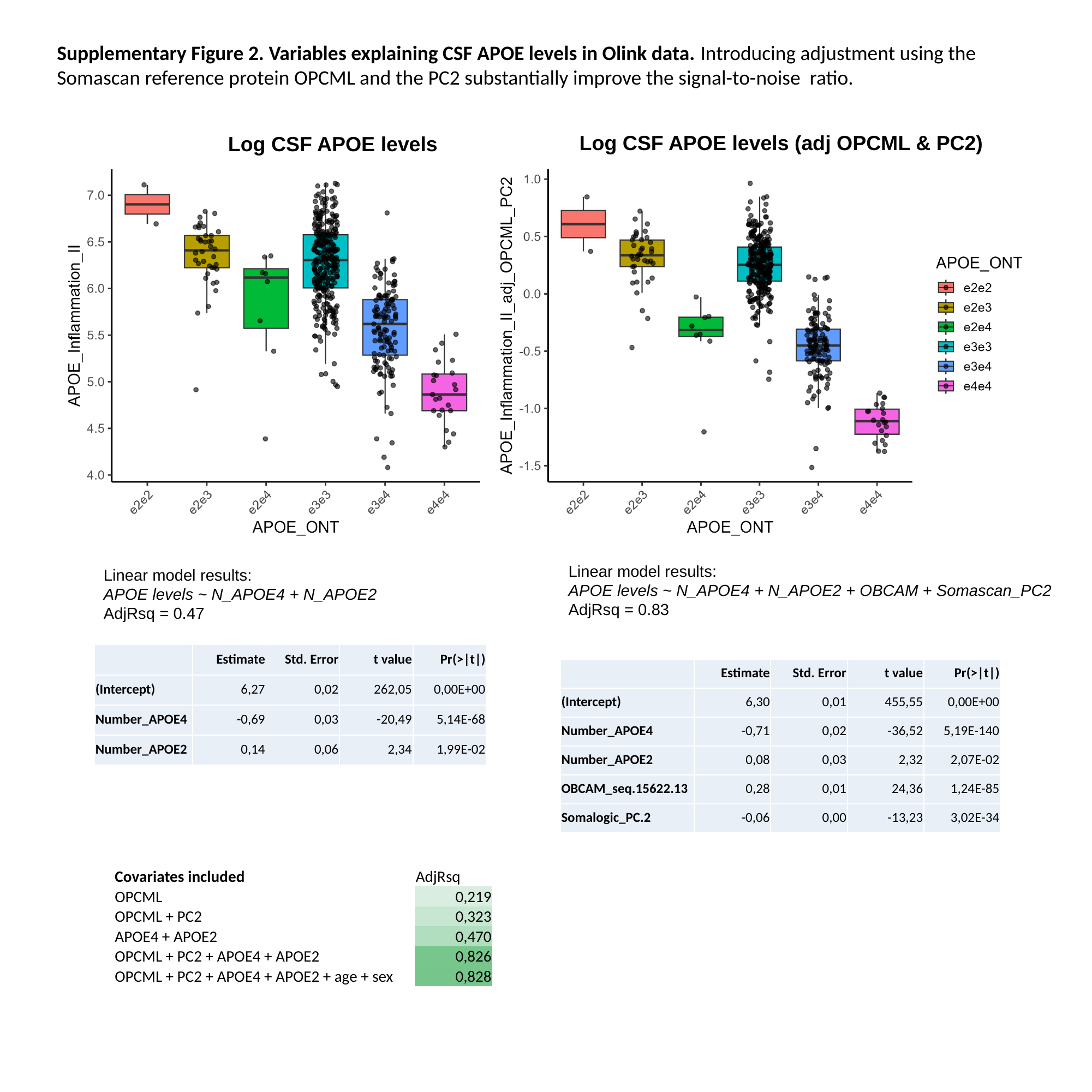

Supplementary Figure 2. Variables explaining CSF APOE levels in Olink data. Introducing adjustment using the Somascan reference protein OPCML and the PC2 substantially improve the signal-to-noise ratio.
Log CSF APOE levels (adj OPCML & PC2)
Log CSF APOE levels
Linear model results:
APOE levels ~ N_APOE4 + N_APOE2 + OBCAM + Somascan_PC2
AdjRsq = 0.83
Linear model results:
APOE levels ~ N_APOE4 + N_APOE2
AdjRsq = 0.47
| | Estimate | Std. Error | t value | Pr(>|t|) |
| --- | --- | --- | --- | --- |
| (Intercept) | 6,27 | 0,02 | 262,05 | 0,00E+00 |
| Number\_APOE4 | -0,69 | 0,03 | -20,49 | 5,14E-68 |
| Number\_APOE2 | 0,14 | 0,06 | 2,34 | 1,99E-02 |
| | Estimate | Std. Error | t value | Pr(>|t|) |
| --- | --- | --- | --- | --- |
| (Intercept) | 6,30 | 0,01 | 455,55 | 0,00E+00 |
| Number\_APOE4 | -0,71 | 0,02 | -36,52 | 5,19E-140 |
| Number\_APOE2 | 0,08 | 0,03 | 2,32 | 2,07E-02 |
| OBCAM\_seq.15622.13 | 0,28 | 0,01 | 24,36 | 1,24E-85 |
| Somalogic\_PC.2 | -0,06 | 0,00 | -13,23 | 3,02E-34 |
| Covariates included | AdjRsq |
| --- | --- |
| OPCML | 0,219 |
| OPCML + PC2 | 0,323 |
| APOE4 + APOE2 | 0,470 |
| OPCML + PC2 + APOE4 + APOE2 | 0,826 |
| OPCML + PC2 + APOE4 + APOE2 + age + sex | 0,828 |

### Slide 3
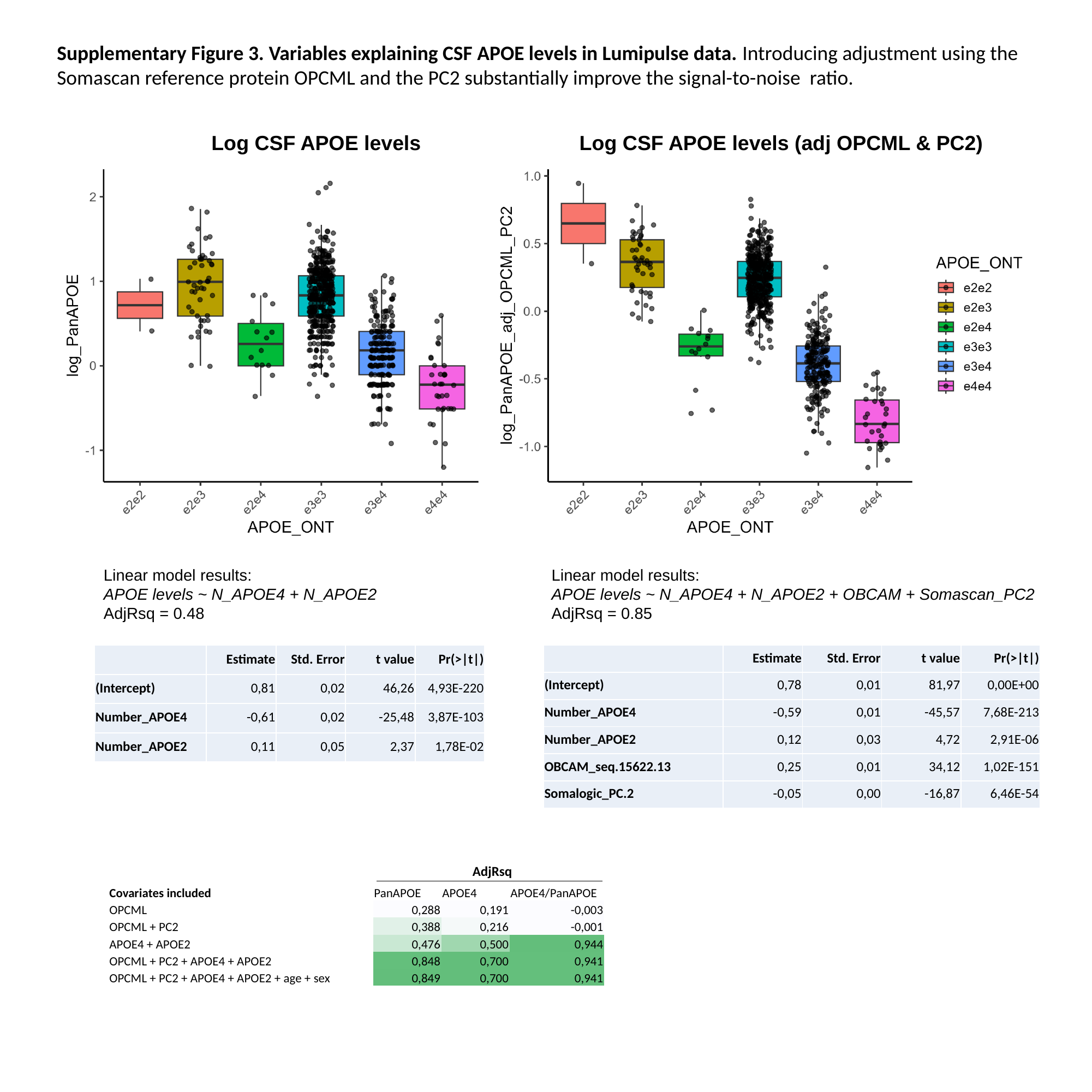

Supplementary Figure 3. Variables explaining CSF APOE levels in Lumipulse data. Introducing adjustment using the Somascan reference protein OPCML and the PC2 substantially improve the signal-to-noise ratio.
Log CSF APOE levels
Log CSF APOE levels (adj OPCML & PC2)
Linear model results:
APOE levels ~ N_APOE4 + N_APOE2
AdjRsq = 0.48
Linear model results:
APOE levels ~ N_APOE4 + N_APOE2 + OBCAM + Somascan_PC2
AdjRsq = 0.85
| | Estimate | Std. Error | t value | Pr(>|t|) |
| --- | --- | --- | --- | --- |
| (Intercept) | 0,78 | 0,01 | 81,97 | 0,00E+00 |
| Number\_APOE4 | -0,59 | 0,01 | -45,57 | 7,68E-213 |
| Number\_APOE2 | 0,12 | 0,03 | 4,72 | 2,91E-06 |
| OBCAM\_seq.15622.13 | 0,25 | 0,01 | 34,12 | 1,02E-151 |
| Somalogic\_PC.2 | -0,05 | 0,00 | -16,87 | 6,46E-54 |
| | Estimate | Std. Error | t value | Pr(>|t|) |
| --- | --- | --- | --- | --- |
| (Intercept) | 0,81 | 0,02 | 46,26 | 4,93E-220 |
| Number\_APOE4 | -0,61 | 0,02 | -25,48 | 3,87E-103 |
| Number\_APOE2 | 0,11 | 0,05 | 2,37 | 1,78E-02 |
AdjRsq
| Covariates included | PanAPOE | APOE4 | APOE4/PanAPOE |
| --- | --- | --- | --- |
| OPCML | 0,288 | 0,191 | -0,003 |
| OPCML + PC2 | 0,388 | 0,216 | -0,001 |
| APOE4 + APOE2 | 0,476 | 0,500 | 0,944 |
| OPCML + PC2 + APOE4 + APOE2 | 0,848 | 0,700 | 0,941 |
| OPCML + PC2 + APOE4 + APOE2 + age + sex | 0,849 | 0,700 | 0,941 |
